# Can Dietary Fibre Intake Reduce the Risk of Mental and Behavioral Disorders Due to Use of Tobacco in Smokers?

**DOI:** 10.64898/2026.03.26.26349460

**Authors:** Xueting Qi, Hui Qi, Na Li, Tong Wang, Weijing Wang, Xin Song, Baibing Mi, Dongfeng Zhang

## Abstract

**Background and aims:** Mental and behavioral disorders due to use of tobacco (MBDT) present a critical challenge to global health, yet modifiable lifestyle factors for reducing its risk remain poorly understood. Given that dietary fibre can affect mental health through gut-brain communication, we sought to explore how fibre intake relates to MBDT risks in smokers.

**Methods:** We specifically evaluated the link between dietary fibre intake and MBDT within a smoking population. Utilizing the UK Biobank (UKB) database, we performed cross-sectional (N=19,943) and prospective cohort (N=19,885) evaluations applying logistic and Cox proportional hazards models, respectively. To determine potential causality, two-sample Mendelian randomization (MR) was applied, relying on GWAS summary data derived from the IEU Open GWAS Project and FinnGen repositories.

**Results:** Cross-sectional findings indicated that individuals in the top quartile (Q4) of fibre intake exhibited decreased MBDT risks relative to the bottom quartile (Q1) (OR: 0.32, 95% CI: 0.13-0.79). Over a median observation time of 12.84 years, the prospective evaluation demonstrated a notable inverse correlation (Q4 HR: 0.46, 95% CI: 0.40-0.54). Non-linear modeling via restricted cubic splines uncovered an L-shaped dose-response curve. Furthermore, MR results confirmed a genetically predicted protective causality (IVW OR: 0.68, 95% CI: 0.49-0.95), which remained consistent across sensitivity validations.

**Conclusions:** Among smokers, higher dietary fibre intake is robustly associated with a reduced risk of mental and behavioral disorders due to the use of tobacco, offering a modifiable dietary target for public health interventions.

## INTRODUCTION

The spectrum of mental and behavioral disorders due to use of tobacco (MBDT)—which includes acute intoxication, withdrawal symptoms, harmful consumption, and dependence—poses a continuous and severe threat to worldwide public health[1–3]. Data from the 2019 Global Burden of Disease study highlight tobacco consumption as a primary driver of global mortality and morbidity, responsible for roughly 7.69 million fatalities and 200 million disability-adjusted life years (DALYs)[1]. Despite comprehensive control measures, the number of smokers remains high, affecting over one billion people worldwide[4]. It is estimated that more than one-third of smokers suffer from mental and behavioral disorders[5]. However, current smoking cessation strategies show poor long-term effectiveness, with relapse rates exceeding 60% to 90% within the first year after attempting to quit[6]. Given this critical bottleneck, identifying modifiable lifestyle factors that could mitigate the risk of MBDT is of paramount importance.

Dietary fibre, a key component of a healthy diet, has long been recognized for its protective effects against cardiometabolic diseases[7]. However, emerging evidence suggests its benefits extend to mental health via the gut-brain axis[8]. Soluble fibre is fermented by gut microbiota to produce short-chain fatty acids (SCFAs), which can modulate systemic inflammation and influence neurotransmitter pathways involved in reward and stress processing[9, 10]. Since neuroinflammation and dopaminergic dysregulation are core features of nicotine addiction, it is biologically plausible that higher dietary fibre intake could confer protection against the development of MBDT[11, 12].

Substantial literature has documented inverse associations between dietary fibre intake and various mental disorders, such as anxiety, psychological distress, and depression[13]. For example, a meta-analysis found that higher dietary fibre intake may offer potential benefits for depressive and anxiety disorders[14]. Likewise, research involving Iranian adults demonstrated an inverse relationship, indicating that greater overall fibre consumption was linked to diminished anxiety and reduced severe psychological distress[15]. Although these findings may imply a potential benefit of dietary fibre on mental health, it remains unclear whether these protective effects extend to smokers. To date, the specific impact of dietary fibre intake on MBDT risk among smokers lacks systematic exploration.

To address these knowledge gaps, this study adopted a comprehensive analytical strategy focusing exclusively on smokers. Initially, we performed a cross-sectional assessment utilizing the UK Biobank (UKB) cohort to explore the baseline link between dietary fibre intake and MBDT among this high-risk population. Subsequently, a cohort study was performed to determine the impact of fibre intake on the long-term risk of developing MBDT during the follow-up period. Finally, to corroborate these observational outcomes and mitigate potential confounding, we applied two-sample Mendelian randomization (MR), relying on summary-level data derived from comprehensive genome-wide association studies.

## METHODS

### Observational study

#### Study design

Established in 2006, the UKB represents one of the most comprehensive biomedical databases worldwide, encompassing health-related data from approximately half a million individuals throughout the United Kingdom[16]. This extensive dataset includes diverse elements such as dietary habits, lifestyle choices, environmental exposures, clinical assessments, genomic profiles, and imaging records. The health status of participants is monitored over time, enabling researchers to investigate longitudinal patterns in disease and wellness.

This research employed both cross-sectional and prospective longitudinal designs. For the baseline analysis, individuals lacking information on smoking history, requisite covariates, or dietary fibre were eliminated. Consequently, the final baseline sample comprised 19,943 individuals. Regarding the prospective cohort, we removed any prevalent cases of MBDT at enrollment, yielding a retained cohort of 19,885 subjects. Each subject’s observation window spanned from their initial recruitment date until December 31, 2022. A detailed schematic outlining these derivation procedures is presented in Supplementary Figure 1.

#### Variable ascertainment

Dietary fibre intake served as the exposure for our analyses. From 2009 to 2010, the UKB utilized the Oxford WebQ to gather participants’ 24-hour dietary recall data based on their food consumption from the previous day[17]. Participants reported their intake using predefined portion sizes across various food groups, thereby improving data precision and consistency[18]. The platform employs integrated food composition databases and computational algorithms to estimate nutrient intake, including dietary fibre intake, automatically[17].

MBDT served as the principal endpoint for our analysis. Case ascertainment was conducted according to the 10th Revision of the International Classification of Diseases (ICD-10), exclusively utilizing the F17 diagnostic category. Comprehensive sub-code specifications are publicly accessible via the UKB data showcase (https://biobank.ndph.ox.ac.uk/showcase/field.cgi?id=41270). Information regarding new cases and their onset times was obtained from hospital admission records.

This study accounted for several covariates, such as age, sex, ethnicity, and others. Definitions and descriptions of these covariates were established based on previously validated study[19].

#### Statistical analysis

For baseline descriptive statistics, continuous variables were expressed as the mean and standard deviation (SD), whereas categorical parameters were detailed using absolute frequencies and relative proportions. Dietary fibre intake was stratified into four equal tiers based on distribution quartiles (Q1 through Q4). The lowest intake group (Q1, <25th percentile) was designated as the baseline reference.

Cross-sectional associations were quantified by estimating odds ratios (ORs) alongside their corresponding 95% confidence intervals (CIs) via logistic regression models. To appraise the longitudinal influence of dietary fibre on MBDT, we computed hazard ratios (HRs) and 95% CIs utilizing Cox proportional hazards models. Observation duration for each subject was recorded from the date of baseline enrollment to whichever came first: the clinical diagnosis of MBDT, mortality, loss to follow-up, or the end of the observation period.

To minimize the influence of potential confounders, we established four progressively adjusted models. Model 1 remained unadjusted. Model 2 incorporated sociodemographic characteristics (age, sex, ethnicity, educational qualifications, employment status, and Townsend deprivation index). Model 3 integrated further adjustments for lifestyle and anthropometric factors, specifically body mass index (BMI), physical activity, alcohol consumption, and energy intake. Finally, Model 4 additionally controlled for baseline cardiometabolic conditions, including diabetes, stroke, and hypertension.

To ensure the reliability of our primary results, we executed multiple sensitivity validations. Initially, to minimize potential reverse causality, we omitted individuals who developed MBDT during the initial 24 months of observation. Second, models were additionally adjusted for sleep duration[20], categorizing 7-8 hours as healthy sleep and all other durations as unhealthy. Third, individuals with extreme dietary fibre or energy intake—defined as below the 1st percentile or above the 99th percentile—were excluded from the analysis. Furthermore, stratified analysis was performed based on sex, age, and BMI, with interaction effects evaluated using the Wald test. Age stratification utilized a 60-year threshold (≤60 vs. >60 years). Additionally, BMI classifications followed standard clinical brackets: <18.5, 18.5 to <25, 25 to <30, and ≥30 kg/m^2^.

We performed linear trend evaluations across intake quartiles to determine if progressively higher fibre intake corresponded to a lowered MBDT susceptibility. Moreover, by treating fibre intake continuously, we utilized restricted cubic splines (RCS) to uncover any potential non-linear dynamics and map the precise trajectory of the dose-response curve.

### Mendelian randomization

#### Study design and data source

Two-sample MR was employed to investigate the causal relationship between dietary fibre and MBDT. We conducted the analysis using the *TwoSampleMR* package version 0.6.17 from R version 4.4.1. The genetic instruments for dietary fibre were obtained from the IEU OpenGWAS project, while summary-level data for MBDT were sourced from the FinnGen consortium. All included studies had received appropriate ethical approvals. Detailed information on the data sources is provided in Supplementary Table 1.

The validity of our MR approach relies on three core tenets: (1) the selected genetic proxies must strongly predict the exposure (relevance assumption); (2) these instruments must not share associations with unmeasured confounders (independence assumption); and (3) the genetic variants must impact the clinical endpoint solely via the primary exposure pathway (exclusion restriction assumption).

#### Selection of genetic instruments

We rigorously filtered the instrumental variables (IVs) by applying several predefined thresholds. Initially, single-nucleotide polymorphisms (SNPs) demonstrating a robust association with dietary fibre intake at a relaxed genome-wide significance threshold (*P*<5×10^-5^) were isolated[21]. Following this, a linkage disequilibrium (LD) clumping procedure (clumping window=10,000 kb, r^2^<0.001) was executed to guarantee the independence of these genetic instruments. Third, to prevent orientation errors, all palindromic variants were systematically removed. Lastly, variants presenting an F-statistic below 10 were discarded to minimize the risk of weak instrument bias.

#### Statistical analysis

We designated the inverse-variance weighted (IVW) as our primary analytical model. To validate the stability of our estimates, we deployed a suite of complementary models, including MR-Egger, weighted median (WM), MR-PRESSO, and Bayesian weighted MR (BWMR). These approaches relax the core assumptions of MR to varying degrees and have been demonstrated to effectively assess the robustness of the findings.

The presence of horizontal pleiotropy was evaluated by calculating the MR-Egger intercept and running the MR-PRESSO global test (*MRPRESSO* package version 1.0). Additionally, inter-instrument heterogeneity was quantified utilizing Cochran’s Q test. Furthermore, we conducted a leave-one-out analysis to ensure the causal inferences were not unduly influenced by any individual SNP. We also executed bidirectional MR to rule out the possibility of reverse causality. Finally, outlier SNPs were identified and excluded using the *RadialMR* package (version 1.1), after which all analyses were re-performed.

The analysis plan for this study was not pre-registered. Therefore, all analyses and findings should be considered exploratory.

## RESULTS

### Observational study results

Baseline demographic and clinical profiles of the 19,943 subjects involved in the cross-sectional evaluation (capturing 58 prevalent cases) are summarized in Table 1. Relative to the lowest intake category (Q1), participants consuming greater amounts of dietary fibre tended to be older males, possess higher educational degrees, and exhibit lower BMI. They also reported elevated energy intake, more vigorous physical activity, and a greater baseline prevalence of diabetes. Within the prospective cohort encompassing 19,885 individuals, we documented 1,819 incident MBDT cases over a median tracking period of 12.84 years.

**Table 1.**
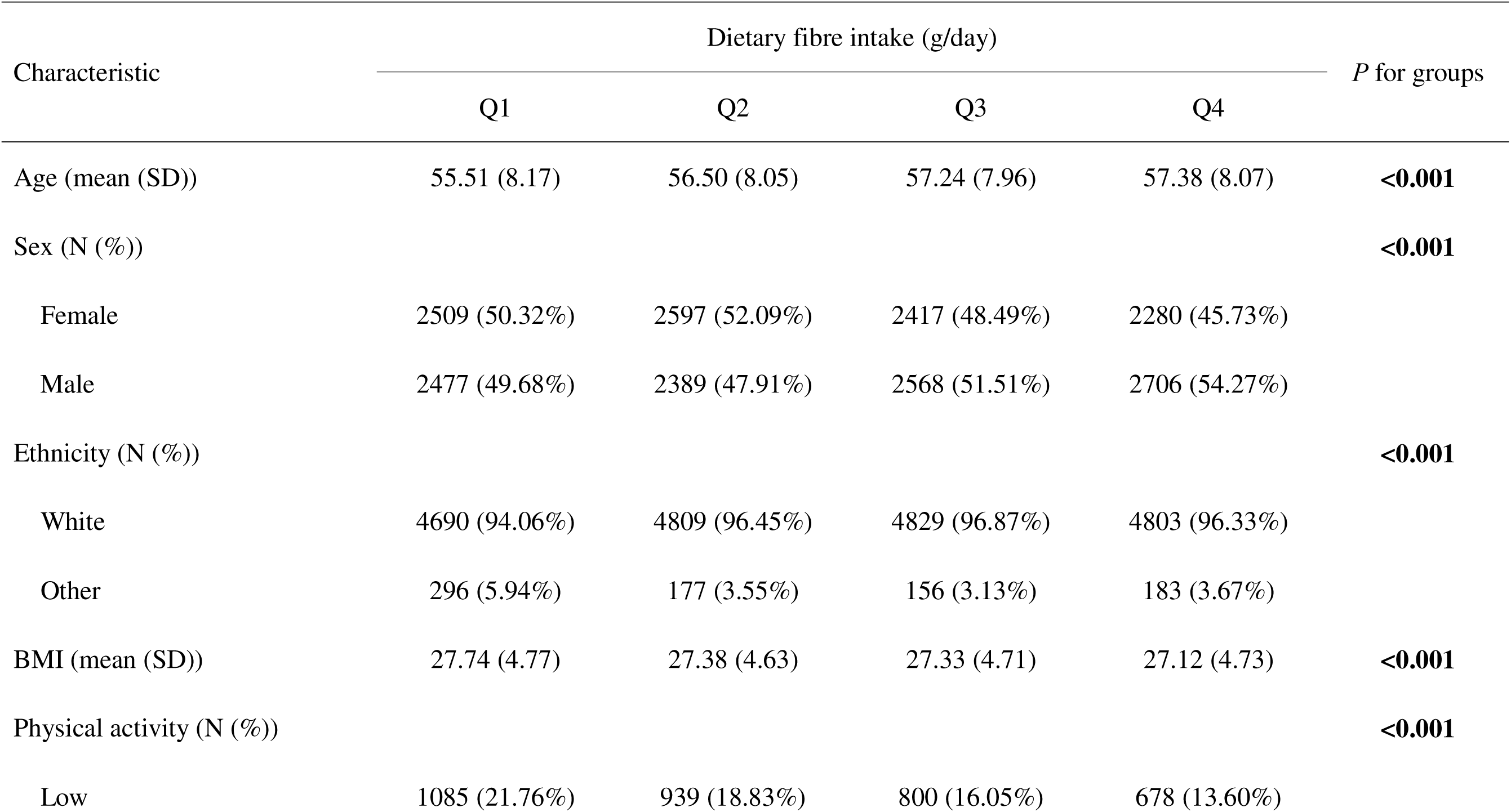
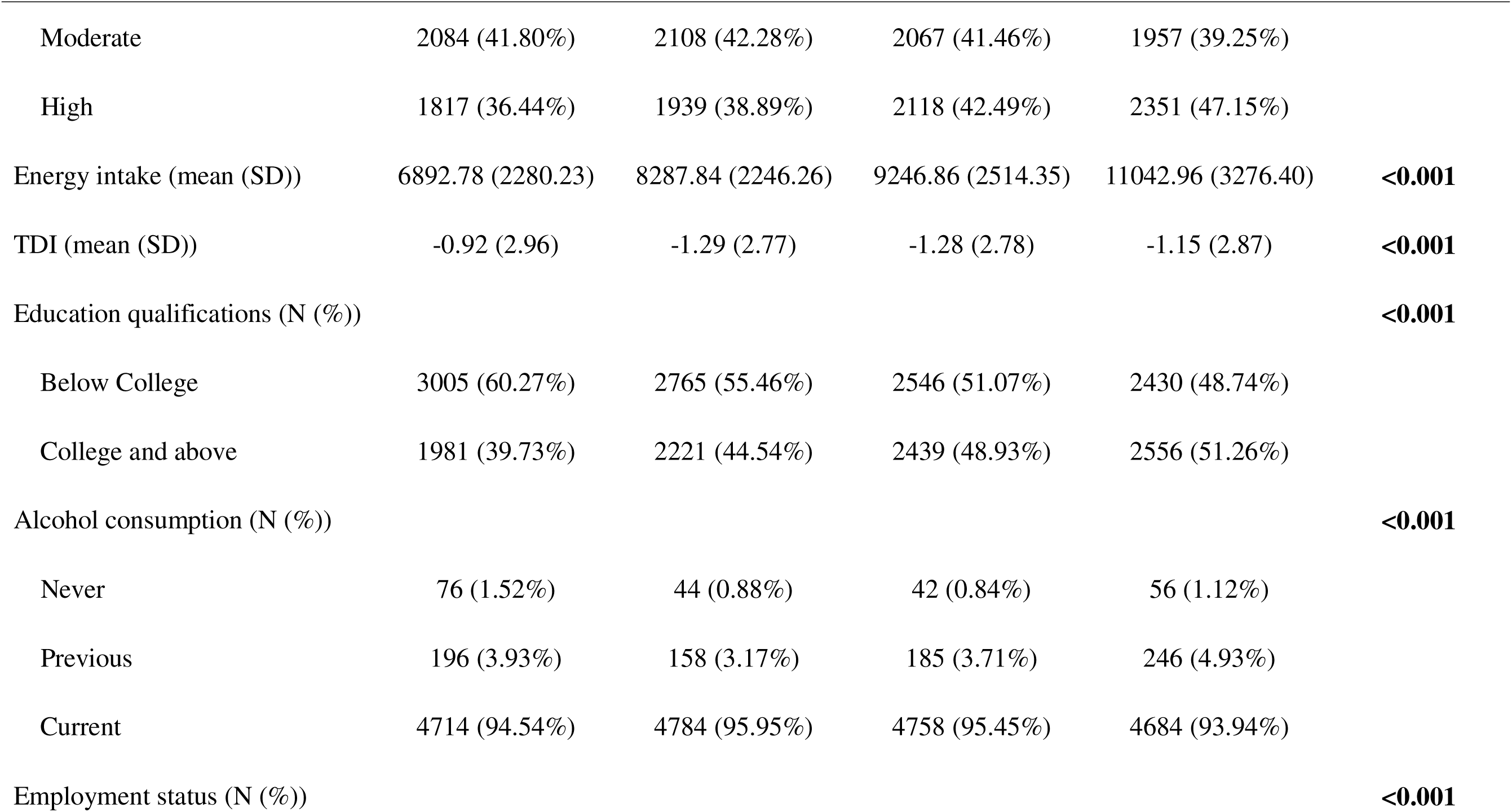
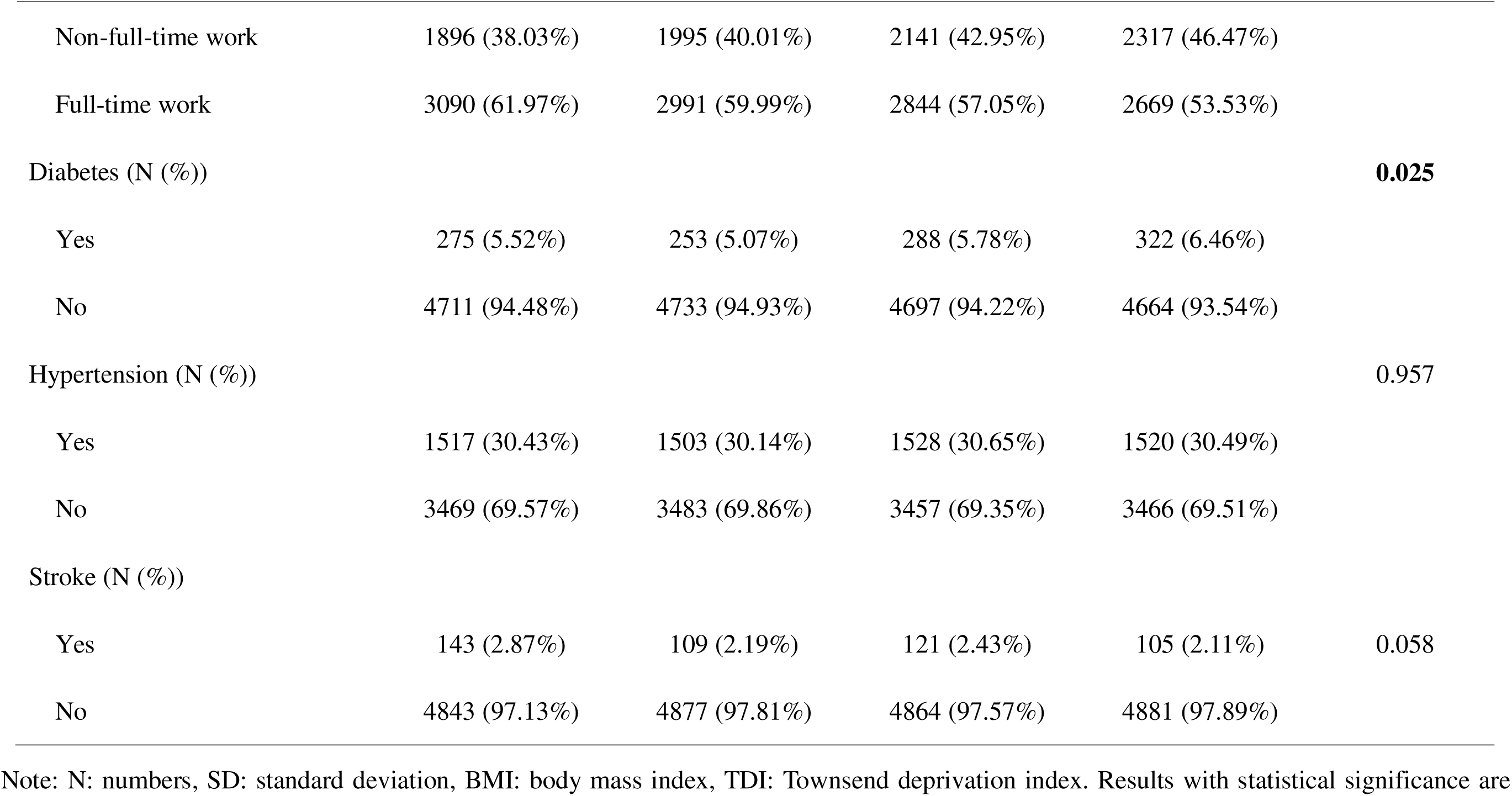
Baseline characteristics of the participants.

Statistical estimates derived from both analytical models are detailed in Table 2. Baseline cross-sectional assessments revealed a pronounced negative correlation between elevated fibre intake and MBDT susceptibility (Q3: OR: 0.39, 95% CI: 0.18-0.86; Q4: OR: 0.32, 95% CI: 0.13-0.79). The proportional hazards assumption was not violated. The results of the cohort study corroborated initial outcomes, confirming a robust protective effect (Q2: HR: 0.62, 95% CI: 0.54-0.70; Q3: HR: 0.53, 95% CI: 0.46-0.61; Q4: HR: 0.46, 95% CI: 0.40-0.54). Importantly, significant dose-response dynamics emerged across both the cross-sectional (*P*=0.008) and longitudinal frameworks (*P*<0.001).

**Table 2.**
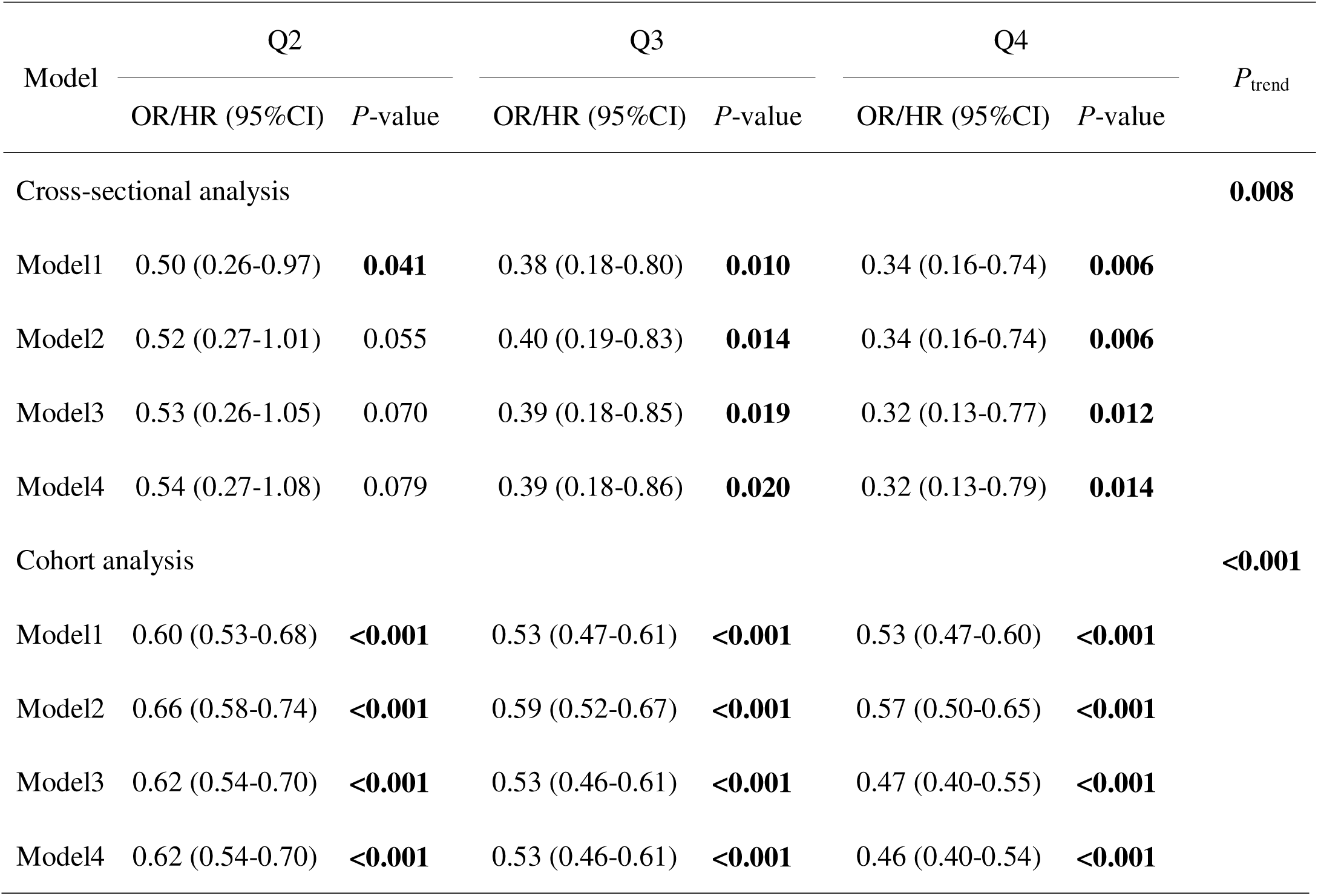
The analysis results of the association between dietary fibre intake and MBDT. Note: OR, odds ratio (reported for the cross-sectional analysis); HR, hazard ratio (reported for the cohort analysis); CI, confidence interval. Model 1 represents the crude, unadjusted estimates. Model 2 incorporates baseline sociodemographic variables (age, sex, ethnic background, educational attainment, occupational status, and Townsend deprivation index). Model 3 additionally controls for lifestyle and physical parameters, specifically BMI, physical activity, total caloric intake, and alcohol consumption. Model 4 introduces further adjustments for underlying cardiometabolic conditions (hypertension, diabetes mellitus, and stroke). The lowest intake quartile (Q1) functioned as the baseline reference category. Results with statistical significance are displayed in bold.

The results of the sensitivity analyses were highly consistent with those of the main analysis (Supplementary Tables 2-4). Following stratification by age and sex, the protective correlations retained high statistical significance (Supplementary Table 5). Conversely, when segmenting the population by BMI, statistically significant risk-reducing effects were observed among normal-weight and overweight participants, though the interaction term was not significant (Supplementary Table 5).

Non-linear modeling utilizing RCS exposed a distinct L-shaped trajectory governing the exposure-outcome link (*P* for nonlinearity<0.001). Specifically, the risk of MBDT decreased precipitously with increasing fibre intake at lower levels, reaching an inflection point at approximately 17.23 g/day. Beyond this threshold, the protective association decelerated and plateaued (Figure 1). Similar associations were observed in stratified analyses by sex and age (Supplementary Figure 2).

**Figure 1.**
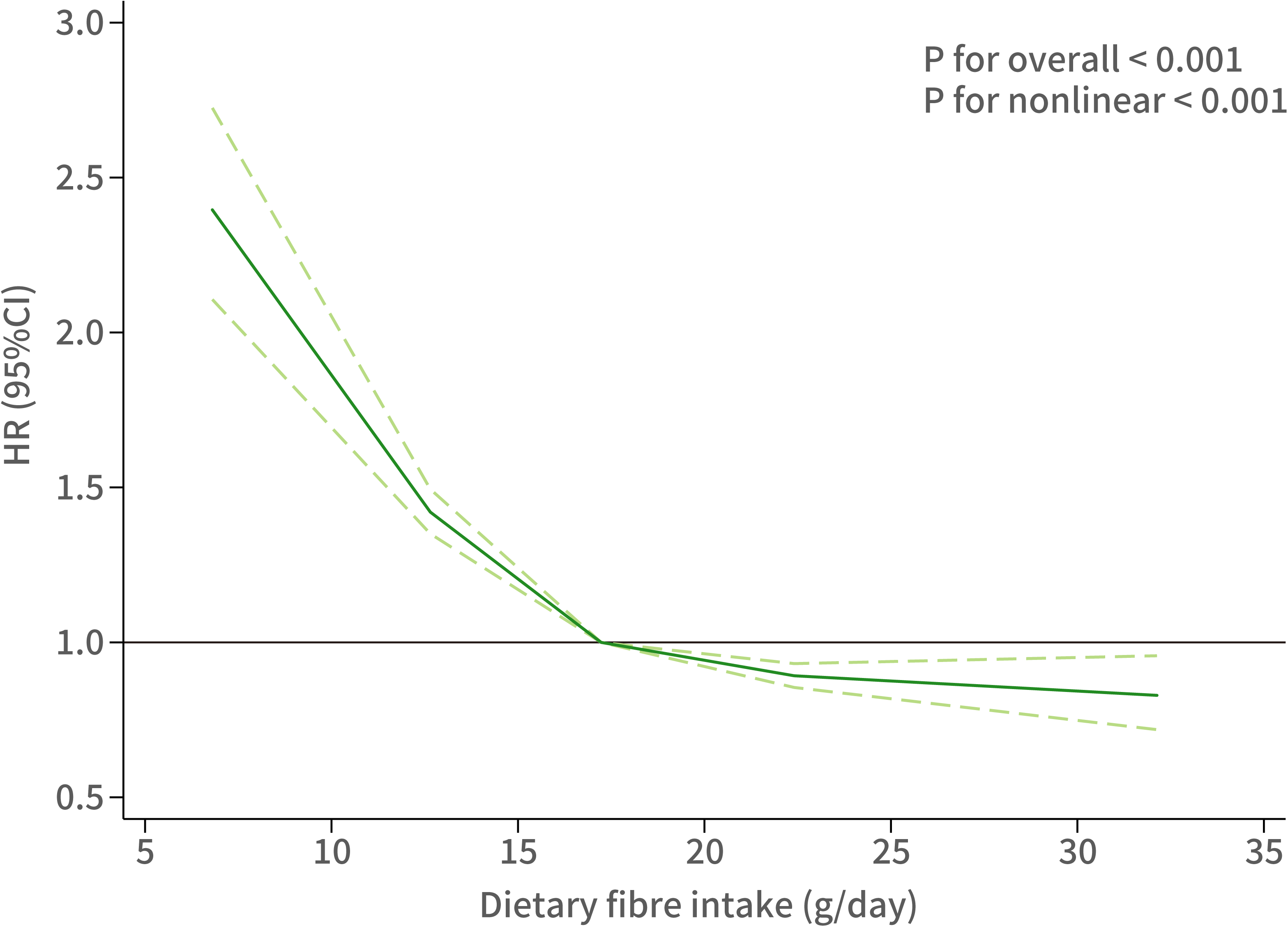
Nonlinear association between dietary fibre intake and MBDT risk Note: HR: hazard ratio; CI: confidence interval.

### Mendelian randomization analysis

The complete set of genetic instruments utilized for the MR framework is detailed in Supplementary Table 6. Primary assessments employing the IVW method provided genetic evidence (OR: 0.68, 95%CI: 0.49-0.95) that augmented fibre intake lowers MBDT susceptibility (Figure 2). This causal inference was further reinforced by highly concordant estimates from both the PRESSO (OR: 0.68, 95%CI: 0.49-0.95) and BWMR (OR: 0.66, 95%CI: 0.47-0.92). No evidence of pleiotropy or heterogeneity bias was detected (Supplementary Tables 7 and 8). Furthermore, leave-one-out validations demonstrated estimate stability, ensuring that causal signals were not disproportionately influenced by any single SNP (Supplementary Figure 3). The results remained significantly unchanged after the removal of outlier SNPs (Supplementary Tables 7-9 and Supplementary Figure 4).

**Figure 2.**
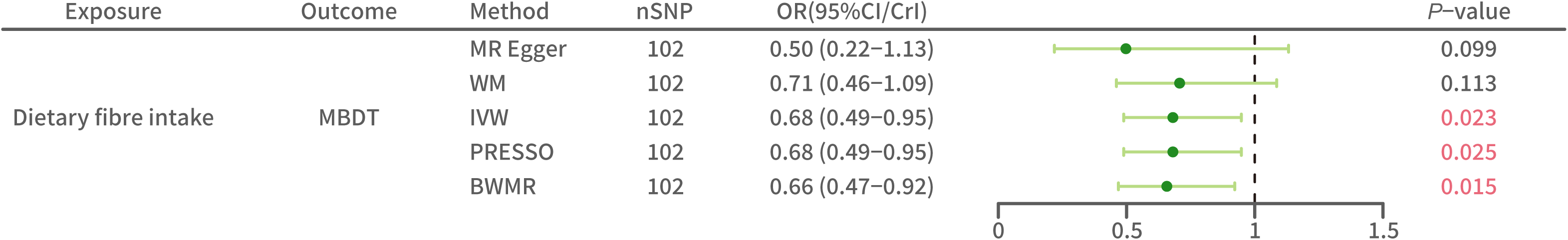
MR result of the association between dietary fibre intake and MBDT Note: SNP: Single nucleotide polymorphism; OR: odds ratio; CI: confidence interval (reported for MR-Egger, WM, IVW, and PRESSO); CrI: credible interval (reported for BWMR); MBDT: mental and behavioral disorders due to the use of tobacco; WM: weighted median; IVW: inverse variance weighted; BWMR: Bayesian weighted Mendelian randomization. Results with statistical significance are displayed in red.

Meanwhile, bidirectional investigations detailed in Supplementary Table 10 lacked clear evidence for a reverse causal effect across all five testing models. These inconclusive findings proved highly stable and independent of pleiotropic interference, heterogeneity, or single-SNP effects. Even following the systematic removal of outlier variants, the evidence regarding a reverse causal link remained inconclusive (Supplementary Tables 11-12 and Supplementary Figures 5-6).

## DISCUSSION

In this large-scale study leveraging data from the UKB and FinnGen consortium, we employed a comprehensive analytical strategy combining cross-sectional, prospective cohort, and MR analyses to determine the precise impact of fibre intake on MBDT susceptibility specifically among smokers. Our observational assessments demonstrated a robust inverse link, revealing that elevated dietary fibre intake corresponds to a decreased vulnerability to MBDT, with RCS analyses revealing a nonlinear dose-response relationship characterized by a steep risk reduction at lower intake levels that stabilizes after a turning point. This pattern suggests a saturation effect, implying that the most substantial benefits are achieved by shifting from low to moderate fibre intake. Crucially, these associations were largely consistent across various sensitivity analyses. Furthermore, genetic evidence from MR analysis provided support for a potential causal protective effect of dietary fibre against MBDT.

While direct evidence linking dietary fibre to MBDTs remains limited, our findings strongly align with an extensive pool of research documenting the mental benefits of fibre consumption[22, 23]. For example, previous cross-sectional analyses utilizing the NHANES database revealed that elevated fibre ingestion correlates with a 20% to 30% decreased probability of developing depression[24]. Another cross-sectional study reported a significant inverse association between fibre intake and anxiety[25]. Furthermore, these results are corroborated by several meta-analyses[26–28]. Beyond common depression and anxiety, dietary fibre also exerts protective effects in Huntington’s disease, which involves complex psychiatric, cognitive, and motor dysfunctions[29]. Collectively, this consistent evidence regarding the neuroprotective role of dietary fibre in mental and behavioral disorders lends additional support for the conclusion of this study.

Although the precise biological pathways linking dietary fibre and MBDT still require further research, this protective effect may mainly be based on the complex multi-organ signaling network known as the gut-brain axis. Prolonged smoking induces widespread toxic effects, with nicotine and volatile agents precipitating oxidative damage. This stress promotes intestinal epithelial apoptosis and suppresses the expression of key tight junction molecules like Occludin and ZO-1[30, 31]. The subsequent increase in intestinal permeability facilitates the systemic influx of lipopolysaccharides, triggering an inflammatory cascade capable of disrupting the blood-brain barrier (BBB)[32, 33]. In contrast, the intake of dietary fibre serves as a vital substrate for anaerobic fermentation by commensal bacteria, significantly increasing the luminal concentration of SCFAs such as butyrate, propionate, and acetate[34]. These SCFAs function as potent histone deacetylase inhibitors and signal through G-protein coupled receptors 41 to reinforce both intestinal and BBB integrity[35–38]. Once within the central nervous system, SCFAs orchestrate neuroimmune responses by driving microglia away from a pro-inflammatory M1 state towards a neuroprotective, anti-inflammatory M2 configuration[39]. Additionally, these gut-derived signals upregulate brain-derived neurotrophic factor (BDNF) and stimulate the PI3K/Akt-mTOR signaling cascade[40]. Such BDNF enhancement is fundamental to maintaining synaptic plasticity and preserving the integrity of the mesolimbic dopaminergic reward circuits, which are typically compromised by chronic nicotine abuse[41, 42].

This study possesses several notable strengths. First, we utilized an integrative analytical framework that combines cross-sectional, prospective cohort, and MR analyses. This approach enables us to reinforce observational findings with genetic instruments, thereby bolstering the reliability of the protective effects while effectively mitigating the impact of unmeasured confounders and reverse causality. Second, the utilization of large-scale, high-quality datasets from the UKB and the FinnGen consortium provided substantial statistical power to explicitly investigate the risk of MBDT among smokers. Third, the consistency of results across various sensitivity analyses and the identification of a nonlinear dose-response relationship through RCS add further depth and robustness to the evidence.

However, some limitations should be acknowledged. First, dietary fibre intake was estimated based on self-reported 24-hour dietary recalls, making the data susceptible to subjective memory lapses and quantification inaccuracies. Nevertheless, dietary habits in adulthood are generally characterized by relative stability. Consistent with this, a validation study within the UKB demonstrated reasonable reproducibility of dietary intakes, suggesting that baseline measurements can serve as a reliable proxy for long-term habitual consumption[43]. Second, while the UKB and GWAS summary statistics is an extensive resource, the participants are predominantly of White European descent, restricting the broader extrapolation of our conclusions to more diverse ancestral populations. Third, the diagnosis of MBDT relied on ICD-10 codes from hospital admission records. This approach might miss individuals with milder symptoms who did not seek hospital-level care, consequently yielding a conservative estimate of the actual clinical burden. Finally, although MR analysis provides evidence for potential causality, it cannot completely rule out the influence of horizontal pleiotropy despite rigorous sensitivity tests.

## CONCLUSION

In summary, this study provides robust observational and genetic evidence supporting a protective role of higher dietary fibre intake against the risk of MBDT among smokers. This study identified a significant inverse association characterized by an L-shaped dose-response relationship, suggesting that even moderate increases in fibre intake may confer substantial benefits for this high-risk population. The findings highlight dietary fibre as a modifiable lifestyle factor that could be integrated into public health strategies to mitigate the risks of MBDT in smokers. Future research, particularly clinical interventions, is warranted to further elucidate the underlying biological mechanisms and confirm the therapeutic efficacy of fibre supplementation in this context.

## AUTHOR CONTRIBUTIONS

**Xueting Qi:** Conceptualization (lead); Data curation (equal); Formal analysis (equal); Methodology (equal); Software (equal); Writing – original draft (lead). **Hui Qi:** Data curation (equal); Formal analysis (equal); Software (equal); Visualization (lead). **Na Li:** Software (equal); Visualization (supporting). **Xin Song:** Visualization (supporting). **Tong Wang:** Resources (equal). **Weijing Wang:** Resources (equal). **Baibing Mi:** Methodology (equal); Writing – review and editing (equal). **Dongfeng Zhang:** Methodology (equal); Writing – review and editing (equal).

## DATA AVAILABILITY STATEMENT

UK Biobank data are available upon successful application (https://www.ukbiobank.ac.uk/enable-your-research/apply-for-access). The GWAS summary statistics used in this study have been publicly released. For dietary iron intake data, please visit https://opengwas.io/datasets/. For the MBDT data, please visit https://www.finngen.fi/en/access_results.

## ACKNOWLEDGEMENT

We sincerely thank all the staff of UKB, IEU Open GWAS Project, and the FinnGen consortium.

## DECLARATION OD INTEREST STATEMENT

The authors declare no competing interests.

## PRIMARY FUNDING

None.

