## Supplementary Material for "Can Dietary Fibre Intake Reduce the Risk of Mental and Behavioral Disorders Due to Use of Tobacco in Smokers?"

**Supplementary Figures Contents**

**Supplementary Tables Contents**

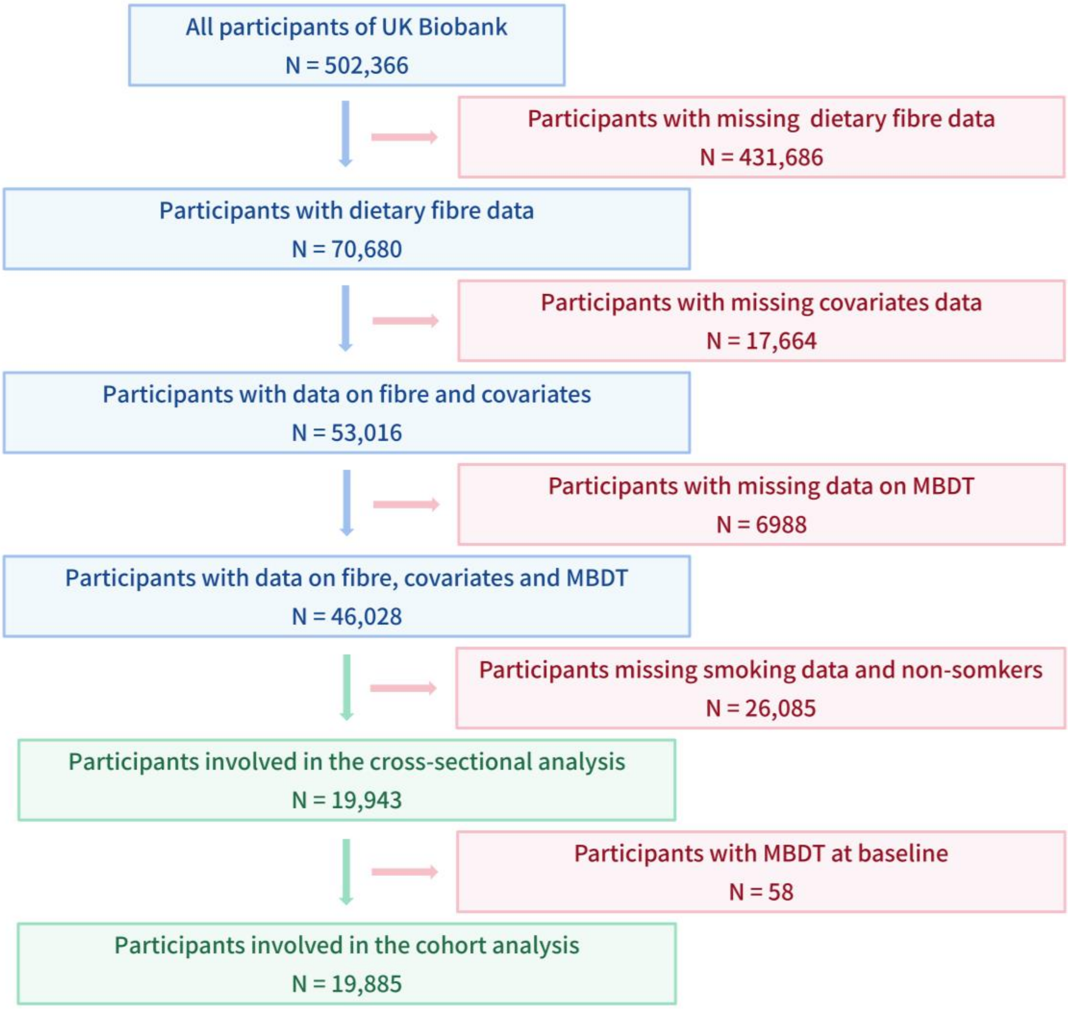

**Supplementary Figure 1** The inclusion process of participants in observational studies

Note: MBDT: mental and behavioral disorders due to use of tobacco.

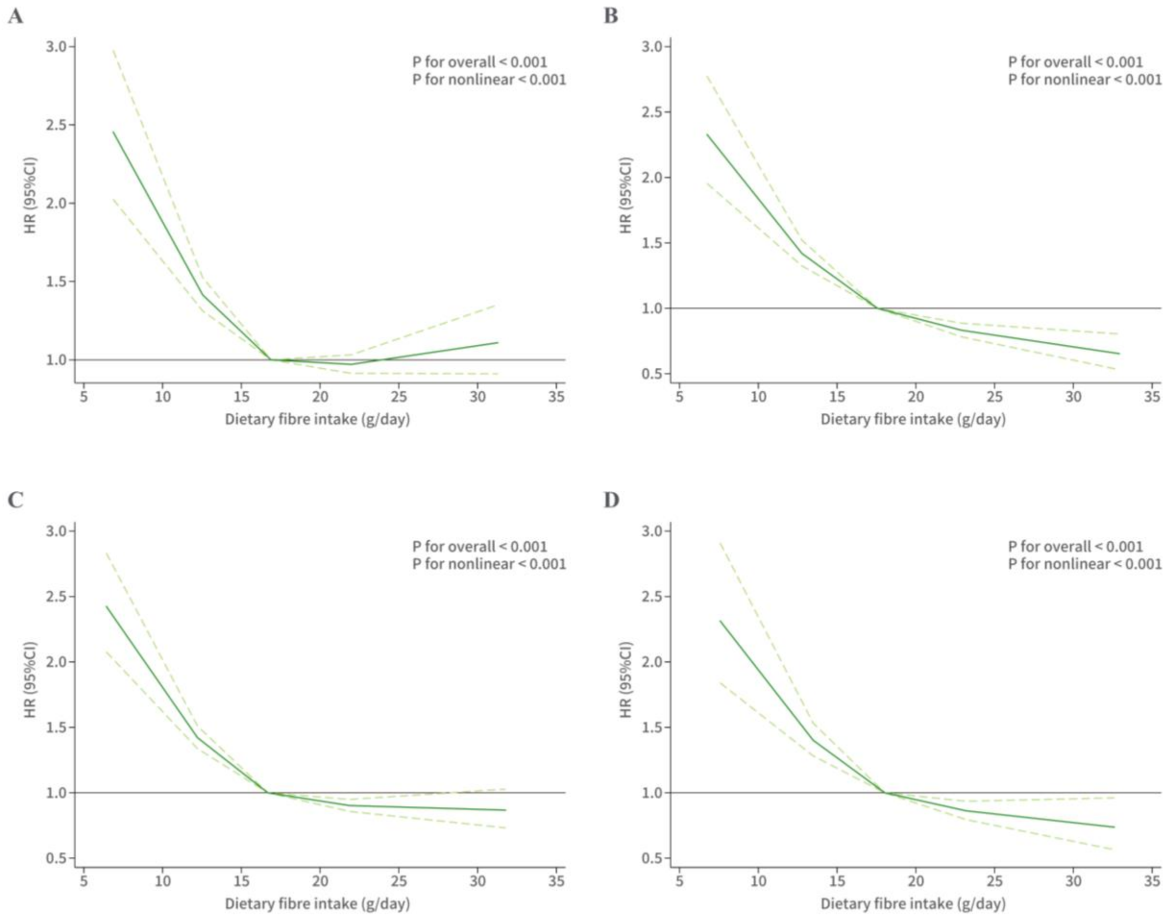

**Supplementary Figure 2** The non-linear association between dietary fiber intake stratified by gender and age and the risk of MBDT

Note: HR: hazard ratio; CI: confidence interval.

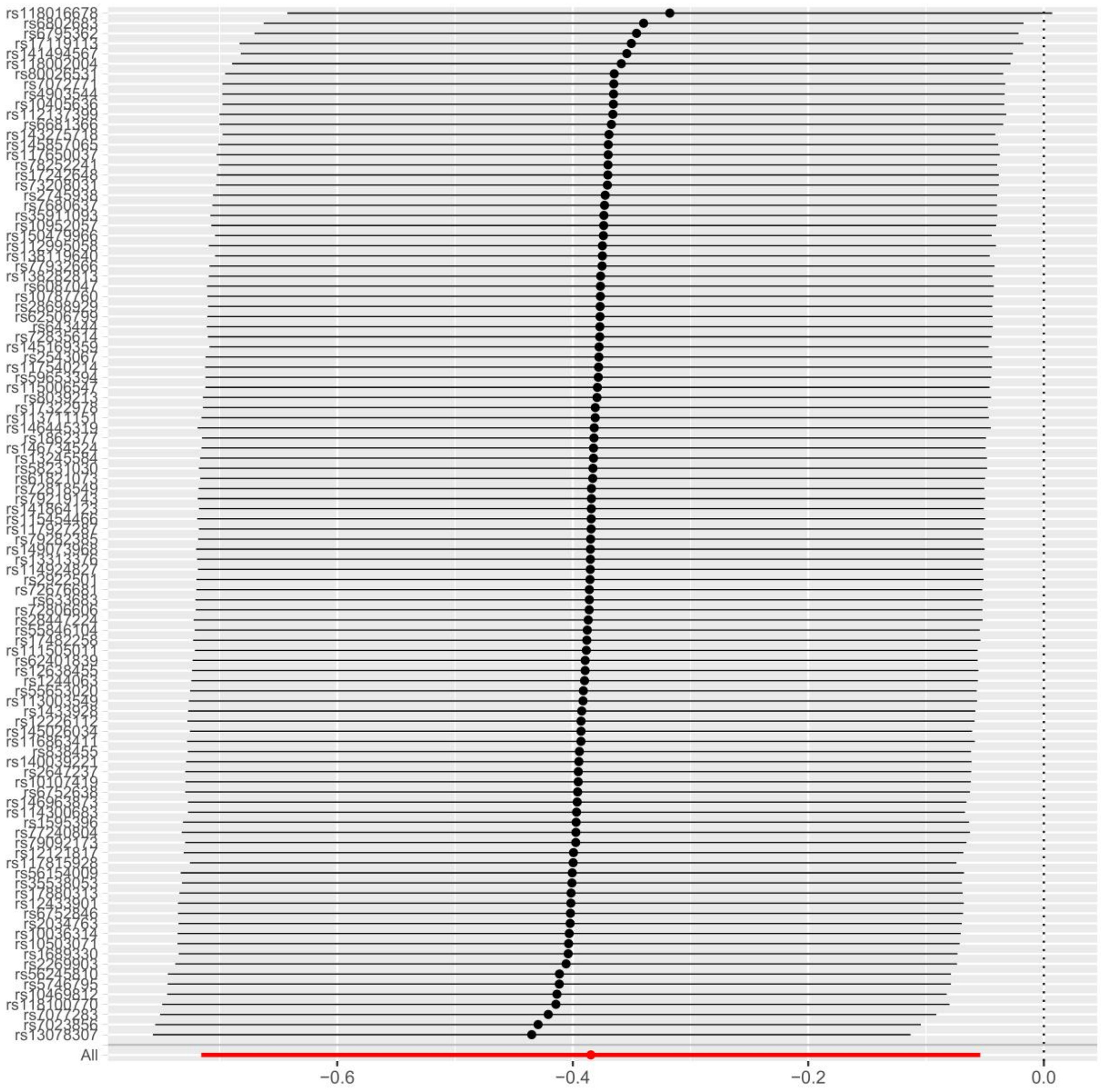

**Supplementary Figure 3** MR leave-one-out analysis for dietary fibre intake on MBDT

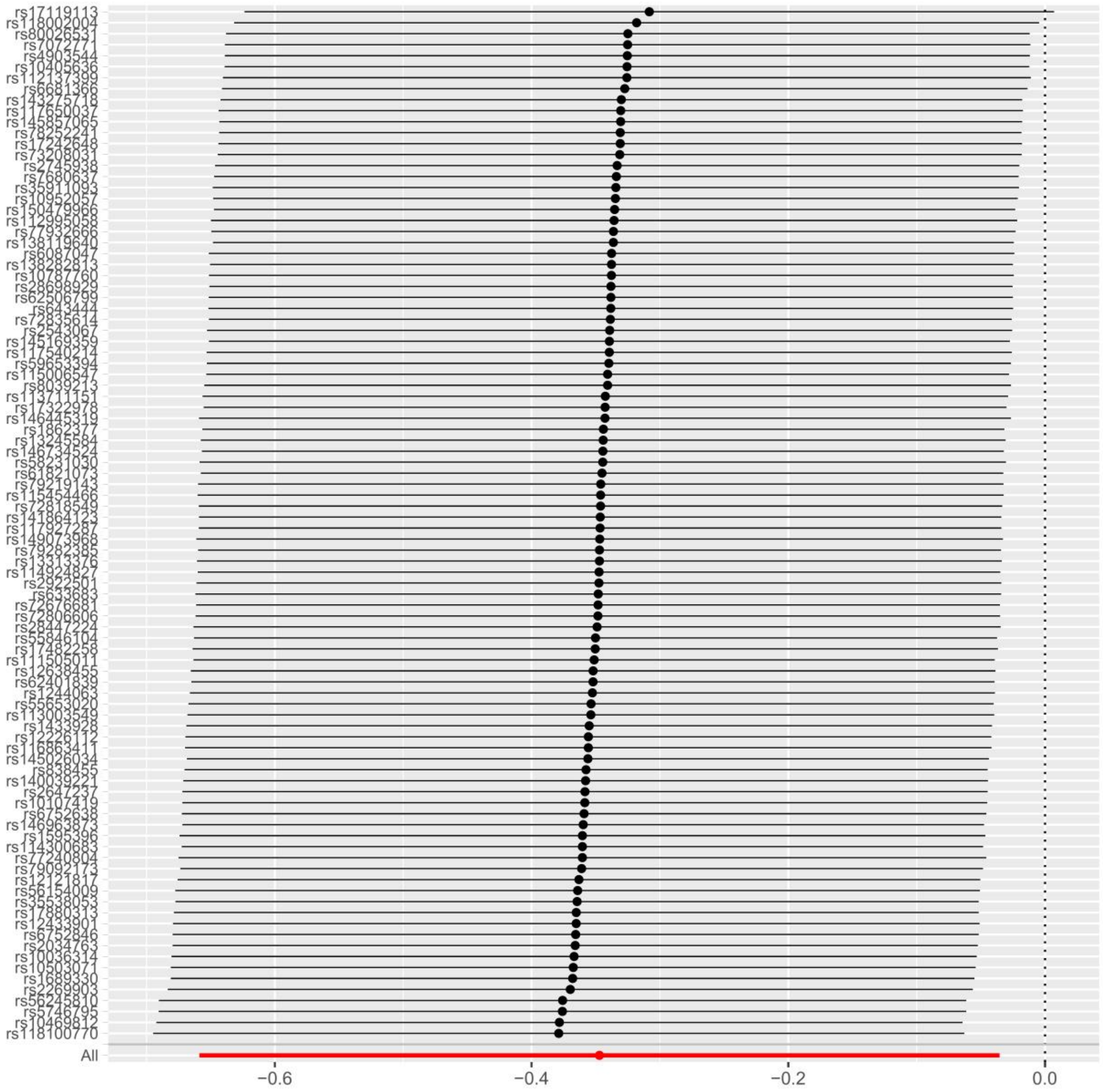

**Supplementary Figure 4** MR leave-one-out analysis for dietary fibre intake on MBDT after excluding outlier SNPs

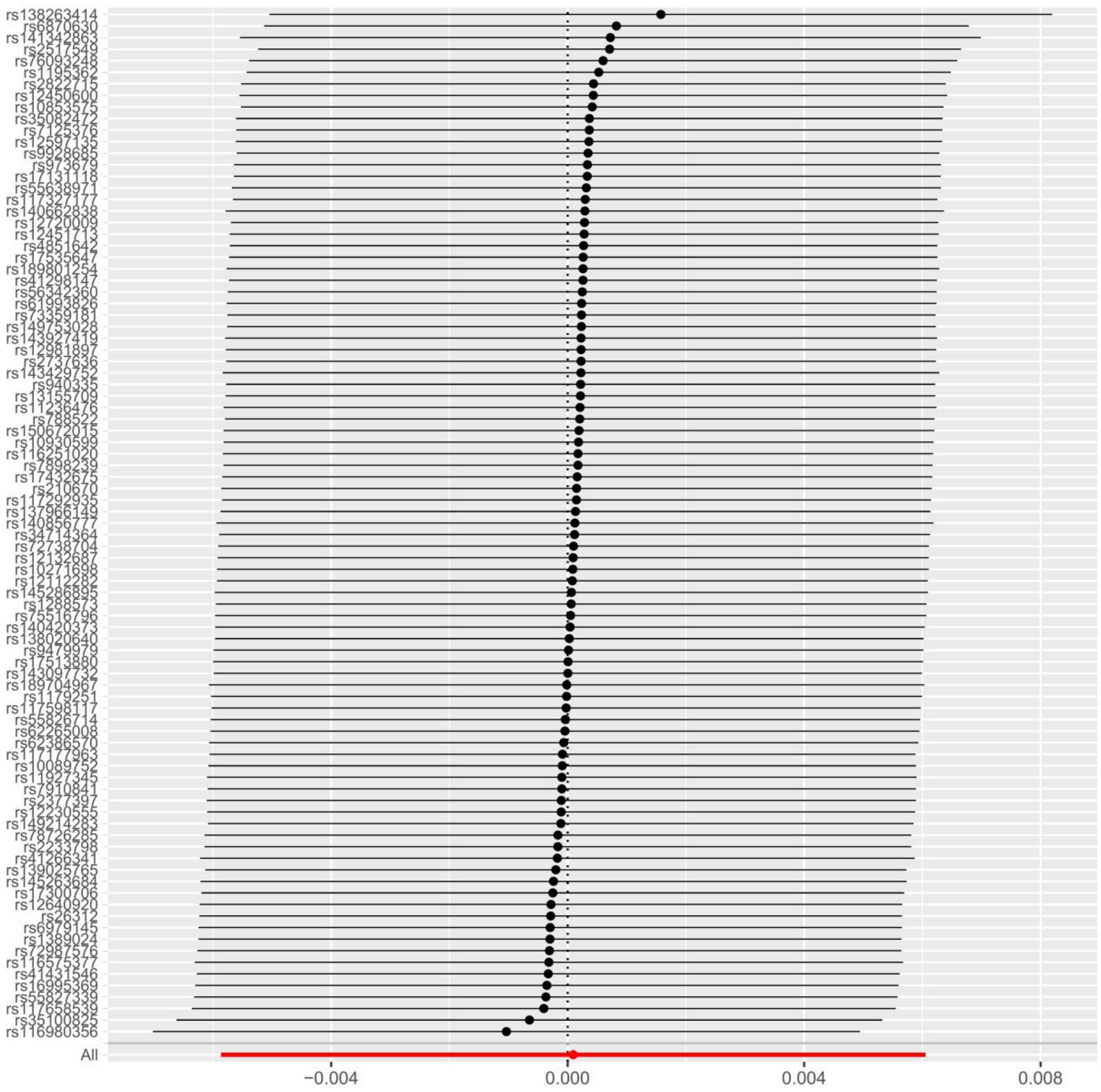

**Supplementary Figure 5** MR leave-one-out analysis for MBDT on dietary fibre intake

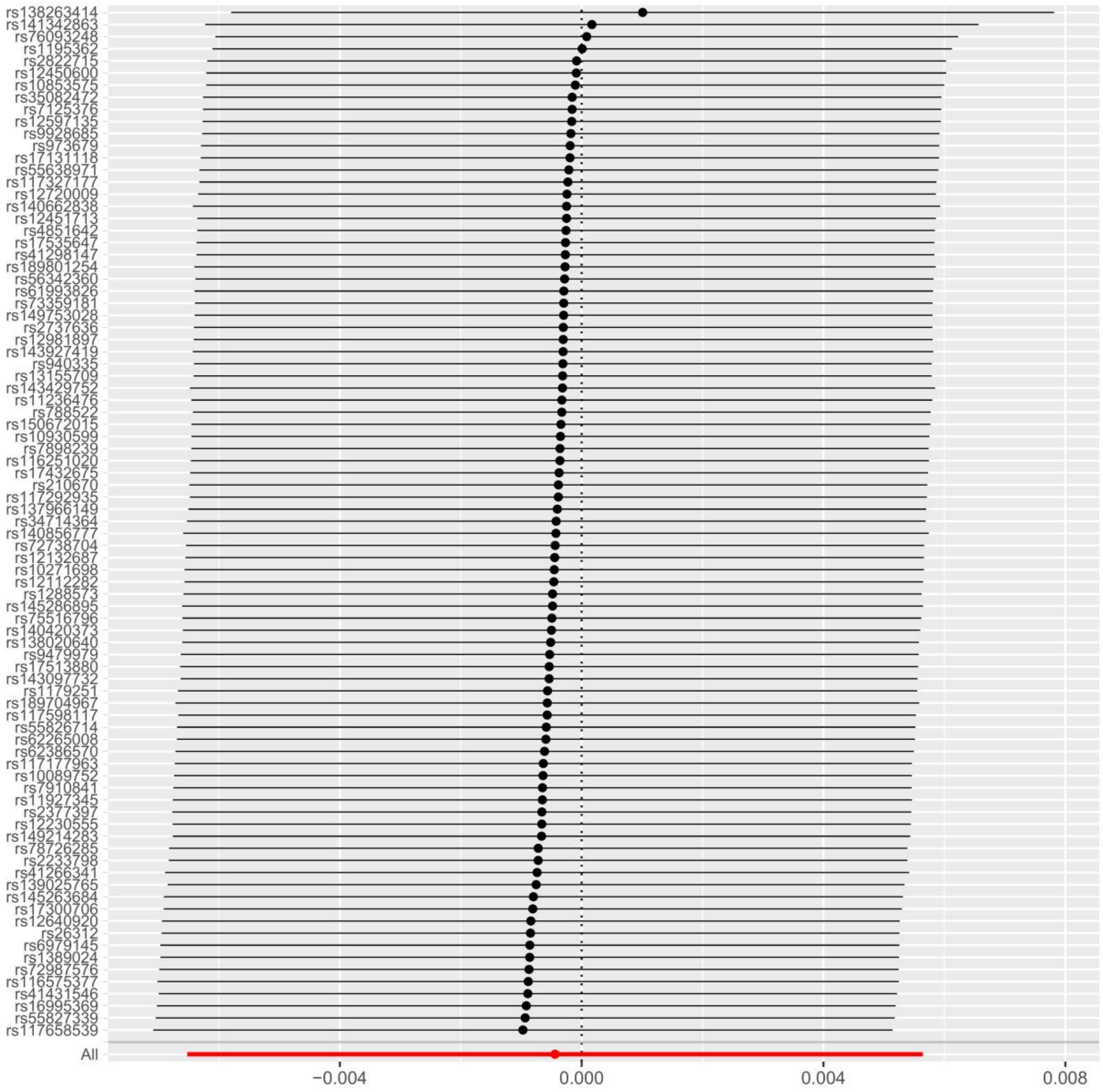

**Supplementary Figure 6** MR leave-one-out analysis for dietary fibre intake on MBDT after excluding outlier SNPs

**Supplementary Table 1** Data source for Mendelian randomization analysis

| Trait | GWAS ID or PMID | Year | Sample size | Ethnicity | URL |
|---|---|---|---|---|---|
| Dietary fibre intake | ukb-b-19085 | 2018 | 64979 | European | https://gwas.mrcieu.ac.uk/datasets/ukb-b-19085/ |
| MBDT | 36653562 | 2022 | 305463 | European | https://storage.googleapis.com/finngen-public-data-r7/summary_stats/finngen_R7_F5_TOBAC.gz |

Note: MBDT: mental and behavioral disorders due to use of tobacco.

**Supplementary Table 2** Analyses result excluding participants diagnosed with MBDT in the two years prior to follow-up

| Model | Q2 | | Q3 | | Q4 | | $P_{trend}$ |
|---|---|---|---|---|---|---|---|
| | HR (95%CI) | *P*-value | HR (95%CI) | *P*-value | HR (95%CI) | *P*-value | |
| Model1 | 0.60 (0.53-0.68) | **<0.001** | 0.54 (0.47-0.62) | **<0.001** | 0.53 (0.46-0.61) | **<0.001** | **<0.001** |
| Model2 | 0.65 (0.57-0.74) | **<0.001** | 0.59 (0.52-0.68) | **<0.001** | 0.57 (0.50-0.66) | **<0.001** | |
| Model3 | 0.61 (0.53-0.70) | **<0.001** | 0.53 (0.46-0.61) | **<0.001** | 0.46 (0.39-0.54) | **<0.001** | |
| Model4 | 0.61 (0.53-0.70) | **<0.001** | 0.53 (0.46-0.61) | **<0.001** | 0.46 (0.39-0.54) | **<0.001** | |

Note: HR: hazard ratio; CI: confidence interval. Model 1 was not adjusted for covariates. Model 2 adjusted for age, sex, ethnicity, education qualifications, employment status, and TDI. Model 3 further adjusted for BMI, physical activity level, energy intake, and alcohol consumption. Model 4 further adjusted for hypertension, diabetes, and stroke. Q1 group was reference group. Results with statistical significance are displayed in bold.

**Supplementary Table 3** Analyses result Analysis results of additional adjustments to sleep duration

| Model | Q2 | | Q3 | | Q4 | | $P_{trend}$ |
|---|---|---|---|---|---|---|---|
| | HR (95%CI) | *P*-value | HR (95%CI) | *P*-value | HR (95%CI) | *P*-value | |
| Model1 | 0.61 (0.54-0.69) | **<0.001** | 0.54 (0.47-0.61) | **<0.001** | 0.53 (0.47-0.61) | **<0.001** | **<0.001** |
| Model2 | 0.66 (0.58-0.75) | **<0.001** | 0.59 (0.52-0.68) | **<0.001** | 0.58 (0.50-0.66) | **<0.001** | |

| Model | Q2 | | Q3 | | Q4 | | $P_{trend}$ |
|---|---|---|---|---|---|---|---|
| | HR (95%CI) | *P*-value | HR (95%CI) | *P*-value | HR (95%CI) | *P*-value | |
| Model3 | 0.62 (0.54-0.70) | **<0.001** | 0.53 (0.47-0.61) | **<0.001** | 0.47 (0.40-0.55) | **<0.001** | |
| Model4 | 0.62 (0.55-0.70) | **<0.001** | 0.53 (0.46-0.61) | **<0.001** | 0.47 (0.40-0.54) | **<0.001** | |

Note: HR: hazard ratio; CI: confidence interval. Model 1 was not adjusted for covariates. Model 2 adjusted for age, sex, ethnicity, education qualifications, employment status, and TDI. Model 3 further adjusted for BMI, physical activity level, energy intake, and alcohol consumption. Model 4 further adjusted for hypertension, diabetes, and stroke. Q1 group was reference group. Results with statistical significance are displayed in bold.

**Supplementary Table 4** Analyses result excluding participants with extreme values of dietary fibre or energy intake

| Model | Q2 | | Q3 | | Q4 | | $P_{trend}$ |
|---|---|---|---|---|---|---|---|
| | HR (95%CI) | *P*-value | HR (95%CI) | *P*-value | HR (95%CI) | *P*-value | |
| Excluding extreme dietary fibre intake | | | | | | | **<0.001** |
| Model1 | 0.64 (0.56-0.72) | **<0.001** | 0.56 (0.49-0.64) | **<0.001** | 0.55 (0.48-0.62) | **<0.001** | |
| Model2 | 0.69 (0.60-0.78) | **<0.001** | 0.62 (0.54-0.70) | **<0.001** | 0.59 (0.51-0.67) | **<0.001** | |
| Model3 | 0.64 (0.56-0.73) | **<0.001** | 0.54 (0.47-0.63) | **<0.001** | 0.47 (0.40-0.55) | **<0.001** | |
| Model4 | 0.64 (0.56-0.73) | **<0.001** | 0.54 (0.47-0.62) | **<0.001** | 0.47 (0.40-0.55) | **<0.001** | |
| Excluding extreme energy intake | | | | | | | **<0.001** |
| Model1 | 0.61 (0.54-0.69) | **<0.001** | 0.54 (0.47-0.62) | **<0.001** | 0.52 (0.46-0.60) | **<0.001** | |
| Model2 | 0.66 (0.58-0.75) | **<0.001** | 0.59 (0.52-0.68) | **<0.001** | 0.57 (0.50-0.65) | **<0.001** | |
| Model3 | 0.62 (0.54-0.70) | **<0.001** | 0.53 (0.46-0.61) | **<0.001** | 0.47 (0.40-0.55) | **<0.001** | |
| Model4 | 0.62 (0.54-0.70) | **<0.001** | 0.53 (0.46-0.61) | **<0.001** | 0.46 (0.40-0.54) | **<0.001** | |

Note: HR: hazard ratio; CI: confidence interval. Model 1 was not adjusted for covariates. Model 2 adjusted for age, sex, ethnicity, education qualifications, employment status, and TDI. Model 3 further adjusted for BMI, physical activity level, energy intake, and alcohol consumption. Model 4 further adjusted for hypertension, diabetes, and stroke. Q1 group was reference group. Results with statistical significance are displayed in bold.

**Supplementary Table 5** Stratified analysis of associations between dietary fibre intake and MBDT based on age, sex, and BMI (Model 4).

| Characters | Q2 | | Q3 | | Q4 | | *P*-value for interaction | $P_{trend}$ |
|---|---|---|---|---|---|---|---|---|
| | HR (95%CI) | *P*-value | HR (95%CI) | *P*-value | HR (95%CI) | *P*-value | | |
| Age | | | | | | | | |
| ≤ 60 years | 0.57 (0.49-0.67) | **<0.001** | 0.52 (0.44-0.62) | **<0.001** | 0.46 (0.38-0.55) | **<0.001** | 0.189 | **<0.001** |
| > 60 years | 0.69 (0.55-0.87) | **0.002** | 0.50 (0.39-0.64) | **<0.001** | 0.44 (0.33-0.58) | **<0.001** | | **<0.001** |
| Sex | | | | | | | | |
| Female | 0.61 (0.51-0.74) | **<0.001** | 0.53 (0.43-0.66) | **<0.001** | 0.55 (0.44-0.70) | **<0.001** | 0.754 | **<0.001** |
| Male | 0.64 (0.53-0.76) | **<0.001** | 0.53 (0.44-0.64) | **<0.001** | 0.41 (0.34-0.51) | **<0.001** | | **<0.001** |
| BMI | | | | | | | | |
| <18.5 kg/m$^2$ | 1.00 (0.17-5.94) | 0.999 | 0.62 (0.07-5.38) | 0.661 | 0.38 (0.03-4.23) | 0.434 | 0.215 | 0.408 |
| ≥18.5 kg/m$^2$ & <25 kg/m$^2$ | 0.49 (0.39-0.61) | **<0.001** | 0.39 (0.30-0.49) | **<0.001** | 0.35 (0.27-0.45) | **<0.001** | | **<0.001** |
| ≥25 kg/m$^2$ & <30 kg/m$^2$ | 0.69 (0.57-0.84) | **<0.001** | 0.55 (0.45-0.68) | **<0.001** | 0.43 (0.33-0.55) | **<0.001** | | **<0.001** |
| ≥30 kg/m$^2$ | 0.66 (0.51-0.86) | **0.002** | 0.73 (0.55-0.95) | **0.022** | 0.76 (0.56-1.02) | 0.070 | | 0.090 |

Note: HR: hazard ratio; CI: confidence interval. Q1 was the reference group. BMI: Body mass index. Model 4 adjusted for age, sex, ethnicity, education qualifications, employment status, TDI, BMI, physical activity level, energy intake, alcohol consumption, hypertension, diabetes, and stroke. Results with statistical significance are displayed in bold.

**Supplementary Table 6** Detailed information of the instrumental variables used for MR analysis

| Trait | SNP | Chr | Position | EA | OA | Beta | Se | EAF | *P*-value | Samplesize | $R^2$ | F | Outlier SNPs |
|---|---|---|---|---|---|---|---|---|---|---|---|---|---|
| Dietary fibre intake | rs10036314 | 5 | 50335164 | A | G | 0.07 | 0.02 | 0.02 | 4.20E-05 | 64979 | 0.00 | 16.77 | FALSE |
| Dietary fibre intake | rs10107419 | 8 | 8439369 | C | T | -0.03 | 0.01 | 0.25 | 3.80E-05 | 64979 | 0.00 | 16.98 | FALSE |
| Dietary fibre intake | rs10405636 | 19 | 18538742 | C | A | -0.02 | 0.01 | 0.35 | 3.20E-05 | 64979 | 0.00 | 17.28 | FALSE |

| Trait | SNP | Chr | Position | EA | OA | Beta | Se | EAF | *P*-value | Samplesize | $R^2$ | F | Outlier SNPs |
|---|---|---|---|---|---|---|---|---|---|---|---|---|---|
| Dietary fibre intake | rs10469812 | 2 | 113948915 | T | C | 0.03 | 0.01 | 0.22 | 1.40E-05 | 64979 | 0.00 | 18.90 | FALSE |
| Dietary fibre intake | rs10503071 | 18 | 60123742 | A | G | 0.04 | 0.01 | 0.10 | 4.20E-05 | 64979 | 0.00 | 16.76 | FALSE |
| Dietary fibre intake | rs10787760 | 10 | 118890693 | A | G | -0.03 | 0.01 | 0.72 | 4.00E-05 | 64979 | 0.00 | 16.86 | FALSE |
| Dietary fibre intake | rs10952057 | 7 | 7435730 | C | T | -0.04 | 0.01 | 0.89 | 3.30E-06 | 64979 | 0.00 | 21.63 | FALSE |
| Dietary fibre intake | rs111505011 | 6 | 45659300 | T | C | -0.09 | 0.02 | 0.02 | 4.70E-05 | 64979 | 0.00 | 16.58 | FALSE |
| Dietary fibre intake | rs112137399 | 2 | 88729291 | T | C | 0.04 | 0.01 | 0.09 | 1.40E-05 | 64979 | 0.00 | 18.80 | FALSE |
| Dietary fibre intake | rs112995058 | 22 | 44069609 | A | G | 0.05 | 0.01 | 0.10 | 4.40E-07 | 64979 | 0.00 | 25.51 | FALSE |
| Dietary fibre intake | rs113003549 | 20 | 60244125 | G | A | -0.05 | 0.01 | 0.07 | 3.80E-05 | 64979 | 0.00 | 16.98 | FALSE |
| Dietary fibre intake | rs113711151 | 5 | 53183644 | T | C | -0.06 | 0.01 | 0.04 | 2.00E-05 | 64979 | 0.00 | 18.16 | FALSE |
| Dietary fibre intake | rs114300683 | 5 | 154885392 | A | G | -0.11 | 0.03 | 0.01 | 5.90E-06 | 64979 | 0.00 | 20.52 | FALSE |
| Dietary fibre intake | rs114924827 | 6 | 470769 | A | G | -0.10 | 0.02 | 0.01 | 4.00E-05 | 64979 | 0.00 | 16.89 | FALSE |
| Dietary fibre intake | rs115006547 | 1 | 94951262 | G | A | 0.10 | 0.02 | 0.01 | 3.00E-05 | 64979 | 0.00 | 17.40 | FALSE |
| Dietary fibre intake | rs115454466 | 3 | 140236499 | T | C | -0.11 | 0.03 | 0.01 | 1.10E-05 | 64979 | 0.00 | 19.36 | FALSE |
| Dietary fibre intake | rs116863411 | 20 | 46633015 | T | A | -0.09 | 0.02 | 0.02 | 1.60E-05 | 64979 | 0.00 | 18.61 | FALSE |
| Dietary fibre intake | rs117540214 | 15 | 84338642 | G | A | -0.05 | 0.01 | 0.06 | 5.60E-06 | 64979 | 0.00 | 20.63 | FALSE |
| Dietary fibre intake | rs117650037 | 15 | 50065224 | A | G | -0.05 | 0.01 | 0.07 | 1.40E-05 | 64979 | 0.00 | 18.87 | FALSE |
| Dietary fibre intake | rs117815928 | 10 | 102843967 | A | G | -0.08 | 0.02 | 0.02 | 1.80E-05 | 64979 | 0.00 | 18.40 | TRUE |
| Dietary fibre intake | rs117927287 | 16 | 20109677 | G | A | 0.09 | 0.02 | 0.02 | 3.60E-05 | 64979 | 0.00 | 17.05 | FALSE |
| Dietary fibre intake | rs118002004 | 11 | 107644423 | C | G | 0.05 | 0.01 | 0.06 | 4.00E-05 | 64979 | 0.00 | 16.88 | FALSE |
| Dietary fibre intake | rs118016678 | 8 | 14329553 | C | G | -0.09 | 0.02 | 0.02 | 3.40E-05 | 64979 | 0.00 | 17.18 | TRUE |
| Dietary fibre intake | rs118100770 | 8 | 122672037 | T | C | 0.16 | 0.03 | 0.01 | 9.80E-07 | 64979 | 0.00 | 23.96 | FALSE |
| Dietary fibre intake | rs12121817 | 1 | 166274431 | G | C | 0.06 | 0.01 | 0.05 | 2.80E-05 | 64979 | 0.00 | 17.53 | FALSE |
| Dietary fibre intake | rs12226112 | 11 | 14163360 | T | G | 0.03 | 0.01 | 0.34 | 4.60E-06 | 64979 | 0.00 | 20.99 | FALSE |

| Trait | SNP | Chr | Position | EA | OA | Beta | Se | EAF | *P*-value | Samplesize | $R^2$ | F | Outlier SNPs |
|---|---|---|---|---|---|---|---|---|---|---|---|---|---|
| Dietary fibre intake | rs12433901 | 14 | 23914573 | C | A | 0.04 | 0.01 | 0.09 | 3.20E-05 | 64979 | 0.00 | 17.32 | FALSE |
| Dietary fibre intake | rs1244063 | 12 | 124832606 | G | A | -0.03 | 0.01 | 0.85 | 1.90E-05 | 64979 | 0.00 | 18.24 | FALSE |
| Dietary fibre intake | rs12638455 | 3 | 25122201 | A | C | 0.02 | 0.01 | 0.58 | 3.90E-05 | 64979 | 0.00 | 16.90 | FALSE |
| Dietary fibre intake | rs13078307 | 3 | 2565355 | A | C | 0.03 | 0.01 | 0.24 | 8.90E-07 | 64979 | 0.00 | 24.15 | TRUE |
| Dietary fibre intake | rs13245584 | 7 | 148990223 | T | C | 0.03 | 0.01 | 0.15 | 2.10E-05 | 64979 | 0.00 | 18.14 | FALSE |
| Dietary fibre intake | rs13313376 | 14 | 63578506 | T | A | -0.03 | 0.01 | 0.15 | 2.20E-05 | 64979 | 0.00 | 18.01 | FALSE |
| Dietary fibre intake | rs138119640 | 1 | 208596034 | T | C | 0.10 | 0.02 | 0.02 | 5.20E-06 | 64979 | 0.00 | 20.76 | FALSE |
| Dietary fibre intake | rs138282813 | 12 | 99493152 | C | G | 0.06 | 0.01 | 0.04 | 4.70E-05 | 64979 | 0.00 | 16.55 | FALSE |
| Dietary fibre intake | rs140039221 | 16 | 57884506 | G | A | -0.05 | 0.01 | 0.06 | 1.40E-05 | 64979 | 0.00 | 18.85 | FALSE |
| Dietary fibre intake | rs141494567 | 5 | 137802002 | A | G | 0.14 | 0.03 | 0.01 | 7.70E-06 | 64979 | 0.00 | 20.00 | TRUE |
| Dietary fibre intake | rs141864123 | 16 | 14755908 | A | G | 0.08 | 0.02 | 0.02 | 1.10E-05 | 64979 | 0.00 | 19.42 | FALSE |
| Dietary fibre intake | rs143275718 | 10 | 97709754 | C | T | 0.10 | 0.03 | 0.01 | 4.60E-05 | 64979 | 0.00 | 16.59 | FALSE |
| Dietary fibre intake | rs1433928 | 18 | 39244966 | C | T | -0.03 | 0.01 | 0.64 | 3.70E-06 | 64979 | 0.00 | 21.41 | FALSE |
| Dietary fibre intake | rs145026034 | 8 | 3075766 | C | T | -0.15 | 0.03 | 0.01 | 9.60E-07 | 64979 | 0.00 | 24.01 | FALSE |
| Dietary fibre intake | rs145169359 | 7 | 41337423 | A | C | 0.10 | 0.02 | 0.02 | 5.60E-06 | 64979 | 0.00 | 20.61 | FALSE |
| Dietary fibre intake | rs145857065 | 5 | 34461430 | C | T | -0.07 | 0.02 | 0.03 | 2.20E-05 | 64979 | 0.00 | 18.04 | FALSE |
| Dietary fibre intake | rs146445319 | 13 | 69307879 | T | C | -0.10 | 0.02 | 0.02 | 3.30E-06 | 64979 | 0.00 | 21.61 | FALSE |
| Dietary fibre intake | rs146734524 | 17 | 35123808 | C | T | 0.10 | 0.02 | 0.02 | 1.40E-05 | 64979 | 0.00 | 18.93 | FALSE |
| Dietary fibre intake | rs146963873 | 16 | 70945259 | A | T | -0.15 | 0.03 | 0.01 | 4.80E-06 | 64979 | 0.00 | 20.93 | FALSE |
| Dietary fibre intake | rs149073968 | 4 | 53749780 | C | G | 0.08 | 0.02 | 0.03 | 2.10E-06 | 64979 | 0.00 | 22.54 | FALSE |
| Dietary fibre intake | rs150479966 | 22 | 31847323 | C | G | 0.16 | 0.03 | 0.01 | 5.20E-06 | 64979 | 0.00 | 20.78 | FALSE |
| Dietary fibre intake | rs1595396 | 2 | 117656927 | T | G | 0.03 | 0.01 | 0.37 | 3.70E-06 | 64979 | 0.00 | 21.41 | FALSE |
| Dietary fibre intake | rs1689330 | 7 | 53102039 | C | G | -0.03 | 0.01 | 0.20 | 8.40E-06 | 64979 | 0.00 | 19.85 | FALSE |

| Trait | SNP | Chr | Position | EA | OA | Beta | Se | EAF | *P*-value | Samplesize | $R^2$ | F | Outlier SNPs |
|---|---|---|---|---|---|---|---|---|---|---|---|---|---|
| Dietary fibre intake | rs17119113 | 1 | 59700843 | G | A | 0.05 | 0.01 | 0.08 | 4.90E-07 | 64979 | 0.00 | 25.32 | FALSE |
| Dietary fibre intake | rs17242648 | 14 | 20898308 | T | C | -0.04 | 0.01 | 0.09 | 4.30E-05 | 64979 | 0.00 | 16.74 | FALSE |
| Dietary fibre intake | rs17322978 | 15 | 70073200 | G | A | -0.04 | 0.01 | 0.08 | 9.90E-06 | 64979 | 0.00 | 19.52 | FALSE |
| Dietary fibre intake | rs17482258 | 12 | 26949117 | T | C | 0.04 | 0.01 | 0.10 | 2.40E-05 | 64979 | 0.00 | 17.87 | FALSE |
| Dietary fibre intake | rs17880313 | 10 | 44880935 | A | G | 0.04 | 0.01 | 0.13 | 8.20E-06 | 64979 | 0.00 | 19.90 | FALSE |
| Dietary fibre intake | rs1862377 | 5 | 160692633 | C | T | 0.12 | 0.03 | 0.01 | 3.10E-05 | 64979 | 0.00 | 17.38 | FALSE |
| Dietary fibre intake | rs2034763 | 9 | 2742608 | T | C | 0.02 | 0.01 | 0.43 | 1.20E-05 | 64979 | 0.00 | 19.12 | FALSE |
| Dietary fibre intake | rs2269903 | 7 | 29247409 | C | A | 0.04 | 0.01 | 0.13 | 3.20E-06 | 64979 | 0.00 | 21.69 | FALSE |
| Dietary fibre intake | rs2543067 | 8 | 39838756 | C | T | 0.03 | 0.01 | 0.72 | 4.70E-06 | 64979 | 0.00 | 20.94 | FALSE |
| Dietary fibre intake | rs2647237 | 4 | 106264589 | C | T | -0.02 | 0.01 | 0.42 | 9.00E-06 | 64979 | 0.00 | 19.72 | FALSE |
| Dietary fibre intake | rs2745938 | 1 | 208139949 | T | G | 0.02 | 0.01 | 0.65 | 2.10E-05 | 64979 | 0.00 | 18.12 | FALSE |
| Dietary fibre intake | rs28447224 | 1 | 924111 | A | T | -0.03 | 0.01 | 0.24 | 6.70E-07 | 64979 | 0.00 | 24.70 | FALSE |
| Dietary fibre intake | rs28698929 | 3 | 178589462 | T | C | -0.05 | 0.01 | 0.06 | 2.30E-05 | 64979 | 0.00 | 17.93 | FALSE |
| Dietary fibre intake | rs2922501 | 8 | 134676921 | A | T | 0.02 | 0.01 | 0.67 | 2.40E-05 | 64979 | 0.00 | 17.84 | FALSE |
| Dietary fibre intake | rs35538053 | 1 | 26826513 | A | T | -0.03 | 0.01 | 0.21 | 4.40E-05 | 64979 | 0.00 | 16.69 | FALSE |
| Dietary fibre intake | rs35911093 | 11 | 1094868 | G | C | -0.04 | 0.01 | 0.09 | 1.30E-05 | 64979 | 0.00 | 19.02 | FALSE |
| Dietary fibre intake | rs4903544 | 14 | 77596398 | T | C | -0.03 | 0.01 | 0.30 | 2.10E-05 | 64979 | 0.00 | 18.14 | FALSE |
| Dietary fibre intake | rs55653020 | 7 | 18975650 | G | A | 0.04 | 0.01 | 0.08 | 1.20E-05 | 64979 | 0.00 | 19.19 | FALSE |
| Dietary fibre intake | rs55846104 | 12 | 90444055 | T | C | 0.03 | 0.01 | 0.14 | 2.50E-05 | 64979 | 0.00 | 17.73 | FALSE |
| Dietary fibre intake | rs56154009 | 5 | 79247901 | C | T | 0.02 | 0.01 | 0.58 | 8.40E-06 | 64979 | 0.00 | 19.83 | FALSE |
| Dietary fibre intake | rs56245810 | 2 | 60358491 | C | T | -0.04 | 0.01 | 0.11 | 1.10E-05 | 64979 | 0.00 | 19.30 | FALSE |
| Dietary fibre intake | rs5746795 | 22 | 19585836 | G | A | -0.03 | 0.01 | 0.31 | 3.60E-07 | 64979 | 0.00 | 25.87 | FALSE |
| Dietary fibre intake | rs58231030 | 3 | 31380236 | T | C | 0.07 | 0.02 | 0.03 | 1.30E-05 | 64979 | 0.00 | 18.94 | FALSE |

| Trait | SNP | Chr | Position | EA | OA | Beta | Se | EAF | *P*-value | Samplesize | $R^2$ | F | Outlier SNPs |
|---|---|---|---|---|---|---|---|---|---|---|---|---|---|
| Dietary fibre intake | rs59653394 | 3 | 72155386 | C | T | 0.03 | 0.01 | 0.22 | 2.70E-05 | 64979 | 0.00 | 17.60 | FALSE |
| Dietary fibre intake | rs6087047 | 20 | 9871771 | C | A | -0.03 | 0.01 | 0.16 | 3.30E-05 | 64979 | 0.00 | 17.23 | FALSE |
| Dietary fibre intake | rs61821073 | 1 | 176362810 | A | G | -0.11 | 0.03 | 0.01 | 3.60E-05 | 64979 | 0.00 | 17.05 | FALSE |
| Dietary fibre intake | rs62401839 | 5 | 143898448 | C | T | 0.05 | 0.01 | 0.05 | 3.10E-05 | 64979 | 0.00 | 17.36 | FALSE |
| Dietary fibre intake | rs62506799 | 8 | 31356250 | C | T | -0.03 | 0.01 | 0.16 | 2.30E-05 | 64979 | 0.00 | 17.92 | FALSE |
| Dietary fibre intake | rs633683 | 11 | 118504742 | C | T | 0.03 | 0.01 | 0.60 | 2.40E-06 | 64979 | 0.00 | 22.27 | FALSE |
| Dietary fibre intake | rs643444 | 18 | 76143140 | G | A | -0.02 | 0.01 | 0.61 | 1.40E-05 | 64979 | 0.00 | 18.80 | FALSE |
| Dietary fibre intake | rs6681366 | 1 | 238433005 | A | G | 0.03 | 0.01 | 0.28 | 2.20E-05 | 64979 | 0.00 | 18.03 | FALSE |
| Dietary fibre intake | rs6752638 | 2 | 191885417 | A | G | 0.02 | 0.01 | 0.38 | 2.50E-05 | 64979 | 0.00 | 17.76 | FALSE |
| Dietary fibre intake | rs6752846 | 2 | 69740091 | T | C | 0.03 | 0.01 | 0.58 | 1.10E-06 | 64979 | 0.00 | 23.69 | FALSE |
| Dietary fibre intake | rs6795362 | 3 | 190817134 | C | T | 0.02 | 0.01 | 0.42 | 4.00E-05 | 64979 | 0.00 | 16.87 | FALSE |
| Dietary fibre intake | rs6802683 | 3 | 191627186 | C | T | -0.02 | 0.01 | 0.37 | 3.70E-05 | 64979 | 0.00 | 17.03 | TRUE |
| Dietary fibre intake | rs7023856 | 9 | 81758804 | A | G | -0.03 | 0.01 | 0.27 | 4.50E-06 | 64979 | 0.00 | 21.02 | TRUE |
| Dietary fibre intake | rs7072771 | 10 | 61524061 | T | C | 0.03 | 0.01 | 0.41 | 7.00E-06 | 64979 | 0.00 | 20.19 | TRUE |
| Dietary fibre intake | rs7077283 | 10 | 12117462 | G | A | 0.03 | 0.01 | 0.13 | 2.90E-05 | 64979 | 0.00 | 17.46 | FALSE |
| Dietary fibre intake | rs72676681 | 1 | 73747975 | T | A | 0.06 | 0.01 | 0.04 | 1.20E-05 | 64979 | 0.00 | 19.11 | TRUE |
| Dietary fibre intake | rs72806606 | 16 | 84459794 | A | G | 0.03 | 0.01 | 0.15 | 1.70E-05 | 64979 | 0.00 | 18.53 | FALSE |
| Dietary fibre intake | rs72818549 | 16 | 88085718 | C | T | 0.03 | 0.01 | 0.14 | 3.40E-05 | 64979 | 0.00 | 17.19 | FALSE |
| Dietary fibre intake | rs72835614 | 10 | 84815387 | A | T | -0.05 | 0.01 | 0.05 | 4.80E-05 | 64979 | 0.00 | 16.51 | FALSE |
| Dietary fibre intake | rs73208031 | 13 | 60941660 | G | C | 0.04 | 0.01 | 0.11 | 2.20E-05 | 64979 | 0.00 | 17.97 | FALSE |
| Dietary fibre intake | rs7680637 | 4 | 82239054 | T | C | -0.02 | 0.01 | 0.35 | 2.70E-05 | 64979 | 0.00 | 17.60 | FALSE |
| Dietary fibre intake | rs77240804 | 6 | 91400155 | T | C | 0.11 | 0.03 | 0.01 | 4.00E-05 | 64979 | 0.00 | 16.89 | FALSE |
| Dietary fibre intake | rs77932666 | 7 | 90072451 | G | A | -0.06 | 0.01 | 0.04 | 5.50E-06 | 64979 | 0.00 | 20.66 | FALSE |

| Trait | SNP | Chr | Position | EA | OA | Beta | Se | EAF | *P*-value | Samplesize | $R^2$ | F | Outlier SNPs |
|---|---|---|---|---|---|---|---|---|---|---|---|---|---|
| Dietary fibre intake | rs78252241 | 9 | 137747447 | A | G | -0.11 | 0.03 | 0.01 | 4.70E-05 | 64979 | 0.00 | 16.58 | FALSE |
| Dietary fibre intake | rs79092173 | 6 | 71898247 | G | A | -0.07 | 0.02 | 0.02 | 4.10E-05 | 64979 | 0.00 | 16.82 | FALSE |
| Dietary fibre intake | rs79219143 | 10 | 75706194 | A | G | 0.07 | 0.01 | 0.05 | 2.80E-06 | 64979 | 0.00 | 21.93 | FALSE |
| Dietary fibre intake | rs79282385 | 8 | 66569750 | T | C | 0.07 | 0.02 | 0.03 | 2.10E-05 | 64979 | 0.00 | 18.11 | FALSE |
| Dietary fibre intake | rs80026531 | 6 | 30744768 | G | A | -0.07 | 0.02 | 0.03 | 1.10E-06 | 64979 | 0.00 | 23.75 | FALSE |
| Dietary fibre intake | rs8039213 | 15 | 98346176 | C | T | 0.03 | 0.01 | 0.34 | 1.50E-06 | 64979 | 0.00 | 23.14 | FALSE |
| Dietary fibre intake | rs838455 | 2 | 232560638 | T | C | -0.04 | 0.01 | 0.08 | 3.60E-05 | 64979 | 0.00 | 17.07 | FALSE |
| MBDT | rs10089752 | 8 | 17023423 | G | A | 0.17 | 0.04 | 0.21 | 3.97E-05 | 305463 | 0.00 | 16.89 | FALSE |
| MBDT | rs10271698 | 7 | 27105060 | A | G | 0.49 | 0.12 | 0.02 | 3.87E-05 | 305463 | 0.00 | 16.93 | FALSE |
| MBDT | rs10853575 | 18 | 49033349 | T | C | 0.15 | 0.04 | 0.36 | 3.00E-05 | 305463 | 0.00 | 17.42 | FALSE |
| MBDT | rs10930599 | 2 | 173476430 | C | T | 0.14 | 0.04 | 0.40 | 4.86E-05 | 305463 | 0.00 | 16.50 | FALSE |
| MBDT | rs11236476 | 11 | 75648130 | T | C | 0.39 | 0.10 | 0.03 | 4.76E-05 | 305463 | 0.00 | 16.54 | FALSE |
| MBDT | rs116251020 | 1 | 156121642 | C | A | -0.54 | 0.13 | 0.03 | 2.99E-05 | 305463 | 0.00 | 17.42 | FALSE |
| MBDT | rs116575377 | 3 | 89590756 | C | T | -0.72 | 0.18 | 0.02 | 4.16E-05 | 305463 | 0.00 | 16.80 | FALSE |
| MBDT | rs116980356 | 11 | 34832338 | C | G | 0.70 | 0.17 | 0.01 | 2.64E-05 | 305463 | 0.00 | 17.66 | TRUE |
| MBDT | rs117177963 | 16 | 89948219 | C | T | 0.66 | 0.15 | 0.01 | 1.54E-05 | 305463 | 0.00 | 18.69 | FALSE |
| MBDT | rs117292935 | 18 | 24424176 | A | G | 0.44 | 0.11 | 0.02 | 4.61E-05 | 305463 | 0.00 | 16.60 | FALSE |
| MBDT | rs117327177 | 16 | 22736907 | C | T | 0.33 | 0.08 | 0.04 | 4.02E-05 | 305463 | 0.00 | 16.86 | FALSE |
| MBDT | rs117598117 | 6 | 129390157 | C | T | -0.34 | 0.08 | 0.07 | 9.18E-06 | 305463 | 0.00 | 19.67 | FALSE |
| MBDT | rs117658539 | 8 | 83573547 | C | T | 1.23 | 0.30 | 0.00 | 3.59E-05 | 305463 | 0.00 | 17.08 | FALSE |
| MBDT | rs1179251 | 12 | 68251271 | C | G | -0.33 | 0.08 | 0.06 | 2.90E-05 | 305463 | 0.00 | 17.48 | FALSE |
| MBDT | rs11927345 | 3 | 147682396 | G | C | -0.31 | 0.07 | 0.07 | 2.36E-05 | 305463 | 0.00 | 17.87 | FALSE |
| MBDT | rs1195362 | 3 | 191106012 | A | G | 0.36 | 0.08 | 0.04 | 1.73E-05 | 305463 | 0.00 | 18.47 | FALSE |

| Trait | SNP | Chr | Position | EA | OA | Beta | Se | EAF | *P*-value | Samplesize | $R^2$ | F | Outlier SNPs |
|---|---|---|---|---|---|---|---|---|---|---|---|---|---|
| MBDT | rs12112282 | 7 | 24358339 | T | A | -0.27 | 0.07 | 0.09 | 4.57E-05 | 305463 | 0.00 | 16.62 | FALSE |
| MBDT | rs12132687 | 1 | 167213059 | A | T | 0.25 | 0.06 | 0.08 | 4.10E-05 | 305463 | 0.00 | 16.83 | FALSE |
| MBDT | rs12230555 | 12 | 4309618 | T | C | -0.15 | 0.04 | 0.55 | 3.41E-05 | 305463 | 0.00 | 17.18 | FALSE |
| MBDT | rs12450600 | 17 | 13570204 | G | A | -0.37 | 0.09 | 0.05 | 4.00E-05 | 305463 | 0.00 | 16.87 | FALSE |
| MBDT | rs12451713 | 17 | 12954257 | G | A | -0.18 | 0.04 | 0.23 | 3.20E-05 | 305463 | 0.00 | 17.30 | FALSE |
| MBDT | rs12597135 | 16 | 85072037 | C | T | 0.15 | 0.04 | 0.29 | 4.34E-05 | 305463 | 0.00 | 16.72 | FALSE |
| MBDT | rs12640920 | 4 | 185838419 | T | C | 0.18 | 0.04 | 0.23 | 7.50E-06 | 305463 | 0.00 | 20.06 | FALSE |
| MBDT | rs12720009 | 7 | 590224 | A | G | -0.15 | 0.04 | 0.66 | 2.89E-05 | 305463 | 0.00 | 17.49 | FALSE |
| MBDT | rs1288573 | 4 | 185325804 | A | G | -0.16 | 0.04 | 0.33 | 3.60E-05 | 305463 | 0.00 | 17.07 | FALSE |
| MBDT | rs12981897 | 19 | 48951030 | C | G | -0.17 | 0.04 | 0.28 | 2.34E-05 | 305463 | 0.00 | 17.89 | FALSE |
| MBDT | rs13155709 | 5 | 172355140 | G | C | -0.16 | 0.04 | 0.28 | 3.71E-05 | 305463 | 0.00 | 17.01 | FALSE |
| MBDT | rs137966149 | 16 | 12151066 | A | C | 0.46 | 0.11 | 0.02 | 2.56E-05 | 305463 | 0.00 | 17.72 | FALSE |
| MBDT | rs138020640 | 10 | 63945602 | C | T | 0.28 | 0.07 | 0.07 | 2.48E-05 | 305463 | 0.00 | 17.78 | FALSE |
| MBDT | rs138263414 | 7 | 74500190 | G | A | -4.15 | 1.00 | 0.00 | 3.22E-05 | 305463 | 0.00 | 17.28 | FALSE |
| MBDT | rs1389024 | 18 | 29663545 | G | T | 0.21 | 0.05 | 0.85 | 3.99E-05 | 305463 | 0.00 | 16.88 | FALSE |
| MBDT | rs139025765 | 4 | 95901198 | G | C | 0.26 | 0.06 | 0.07 | 3.95E-05 | 305463 | 0.00 | 16.89 | FALSE |
| MBDT | rs140420373 | 4 | 112968885 | T | C | 0.53 | 0.13 | 0.01 | 4.09E-05 | 305463 | 0.00 | 16.83 | FALSE |
| MBDT | rs140662838 | 3 | 87515674 | A | G | -1.48 | 0.35 | 0.01 | 2.07E-05 | 305463 | 0.00 | 18.12 | FALSE |
| MBDT | rs140856777 | 16 | 29199037 | G | A | -1.00 | 0.24 | 0.01 | 4.27E-05 | 305463 | 0.00 | 16.75 | FALSE |
| MBDT | rs141342863 | 22 | 47651468 | C | T | -2.74 | 0.66 | 0.00 | 3.44E-05 | 305463 | 0.00 | 17.16 | FALSE |
| MBDT | rs143097732 | 1 | 89168548 | C | A | 0.44 | 0.10 | 0.02 | 1.54E-05 | 305463 | 0.00 | 18.69 | FALSE |
| MBDT | rs143429752 | 21 | 27655111 | C | T | -1.23 | 0.30 | 0.01 | 4.88E-05 | 305463 | 0.00 | 16.49 | FALSE |
| MBDT | rs143927419 | 9 | 126100146 | G | A | 0.96 | 0.23 | 0.00 | 3.93E-05 | 305463 | 0.00 | 16.91 | FALSE |

| Trait | SNP | Chr | Position | EA | OA | Beta | Se | EAF | *P*-value | Samplesize | $R^2$ | F | Outlier SNPs |
|---|---|---|---|---|---|---|---|---|---|---|---|---|---|
| MBDT | rs145263684 | 9 | 102129461 | G | A | 0.54 | 0.13 | 0.01 | 2.25E-05 | 305463 | 0.00 | 17.97 | FALSE |
| MBDT | rs145286895 | 2 | 15810109 | C | G | 1.28 | 0.31 | 0.00 | 3.69E-05 | 305463 | 0.00 | 17.02 | FALSE |
| MBDT | rs149214283 | 3 | 180809185 | C | T | 0.61 | 0.14 | 0.01 | 2.16E-05 | 305463 | 0.00 | 18.04 | FALSE |
| MBDT | rs149753028 | 6 | 65624141 | T | C | 0.62 | 0.15 | 0.01 | 3.84E-05 | 305463 | 0.00 | 16.95 | FALSE |
| MBDT | rs150672015 | 8 | 30901871 | T | A | -0.53 | 0.13 | 0.03 | 2.09E-05 | 305463 | 0.00 | 18.11 | FALSE |
| MBDT | rs16995369 | 22 | 35140523 | A | G | -0.24 | 0.06 | 0.11 | 3.01E-05 | 305463 | 0.00 | 17.41 | FALSE |
| MBDT | rs17131118 | 1 | 90646221 | C | T | 0.27 | 0.07 | 0.06 | 4.67E-05 | 305463 | 0.00 | 16.58 | FALSE |
| MBDT | rs17300706 | 13 | 89253947 | A | T | 0.46 | 0.11 | 0.02 | 3.77E-05 | 305463 | 0.00 | 16.98 | FALSE |
| MBDT | rs17432675 | 1 | 201918593 | T | C | 0.15 | 0.04 | 0.36 | 2.39E-05 | 305463 | 0.00 | 17.85 | FALSE |
| MBDT | rs17513880 | 3 | 112374566 | G | A | 0.20 | 0.05 | 0.13 | 4.82E-05 | 305463 | 0.00 | 16.52 | FALSE |
| MBDT | rs17535647 | 4 | 45848903 | T | A | 0.20 | 0.05 | 0.13 | 4.58E-05 | 305463 | 0.00 | 16.61 | FALSE |
| MBDT | rs189704967 | 7 | 90334385 | C | A | 1.34 | 0.32 | 0.00 | 2.69E-05 | 305463 | 0.00 | 17.63 | FALSE |
| MBDT | rs189801254 | 10 | 66576479 | C | A | 0.91 | 0.20 | 0.00 | 7.38E-06 | 305463 | 0.00 | 20.09 | FALSE |
| MBDT | rs210670 | 8 | 40321577 | T | C | -0.16 | 0.04 | 0.70 | 3.46E-05 | 305463 | 0.00 | 17.15 | FALSE |
| MBDT | rs2233798 | 9 | 19048959 | C | T | -0.16 | 0.04 | 0.35 | 2.17E-05 | 305463 | 0.00 | 18.04 | FALSE |
| MBDT | rs2377397 | 17 | 79302071 | T | C | -0.18 | 0.04 | 0.30 | 6.33E-06 | 305463 | 0.00 | 20.39 | FALSE |
| MBDT | rs2517549 | 6 | 31040821 | C | A | 0.16 | 0.04 | 0.27 | 2.73E-05 | 305463 | 0.00 | 17.60 | TRUE |
| MBDT | rs26312 | 3 | 10291174 | G | A | 0.21 | 0.05 | 0.14 | 1.50E-05 | 305463 | 0.00 | 18.73 | FALSE |
| MBDT | rs2737636 | 1 | 200043995 | C | A | -0.27 | 0.07 | 0.09 | 3.23E-05 | 305463 | 0.00 | 17.28 | FALSE |
| MBDT | rs2822715 | 21 | 14517899 | T | C | 0.17 | 0.04 | 0.40 | 2.78E-06 | 305463 | 0.00 | 21.96 | FALSE |
| MBDT | rs34714364 | 1 | 150262258 | G | T | -0.21 | 0.05 | 0.14 | 5.00E-05 | 305463 | 0.00 | 16.45 | FALSE |
| MBDT | rs35082472 | 2 | 17099000 | G | A | 0.18 | 0.05 | 0.17 | 4.85E-05 | 305463 | 0.00 | 16.50 | FALSE |
| MBDT | rs35100825 | 1 | 17651674 | C | A | 0.59 | 0.14 | 0.01 | 3.48E-05 | 305463 | 0.00 | 17.14 | TRUE |

| Trait | SNP | Chr | Position | EA | OA | Beta | Se | EAF | *P*-value | Samplesize | $R^2$ | F | Outlier SNPs |
|---|---|---|---|---|---|---|---|---|---|---|---|---|---|
| MBDT | rs41266341 | 6 | 158585341 | A | C | 1.45 | 0.36 | 0.00 | 4.88E-05 | 305463 | 0.00 | 16.50 | FALSE |
| MBDT | rs41298147 | 9 | 137080953 | G | A | -0.22 | 0.05 | 0.18 | 3.30E-06 | 305463 | 0.00 | 21.63 | FALSE |
| MBDT | rs41431546 | 1 | 215984990 | A | C | 0.17 | 0.04 | 0.27 | 1.48E-05 | 305463 | 0.00 | 18.76 | FALSE |
| MBDT | rs4851642 | 2 | 102942933 | C | G | 0.21 | 0.05 | 0.13 | 2.25E-05 | 305463 | 0.00 | 17.96 | FALSE |
| MBDT | rs55638971 | 2 | 78299530 | T | G | 0.25 | 0.05 | 0.11 | 2.02E-06 | 305463 | 0.00 | 22.57 | FALSE |
| MBDT | rs55826714 | 14 | 43748255 | C | T | 0.22 | 0.05 | 0.11 | 4.04E-05 | 305463 | 0.00 | 16.85 | FALSE |
| MBDT | rs55827339 | 7 | 36693795 | T | C | 0.21 | 0.05 | 0.14 | 1.06E-05 | 305463 | 0.00 | 19.41 | FALSE |
| MBDT | rs56342360 | 15 | 94009044 | C | T | -0.14 | 0.03 | 0.53 | 3.35E-05 | 305463 | 0.00 | 17.21 | FALSE |
| MBDT | rs61993826 | 15 | 25908174 | T | C | 0.36 | 0.09 | 0.03 | 2.84E-05 | 305463 | 0.00 | 17.52 | FALSE |
| MBDT | rs62265008 | 3 | 90387084 | C | T | 0.35 | 0.08 | 0.04 | 3.26E-05 | 305463 | 0.00 | 17.26 | FALSE |
| MBDT | rs62386570 | 6 | 4785409 | A | G | -0.29 | 0.07 | 0.09 | 2.43E-05 | 305463 | 0.00 | 17.82 | FALSE |
| MBDT | rs6870630 | 5 | 26437101 | T | C | 0.35 | 0.08 | 0.95 | 4.01E-05 | 305463 | 0.00 | 16.87 | TRUE |
| MBDT | rs6979145 | 7 | 136133727 | C | G | 0.27 | 0.06 | 0.08 | 8.11E-06 | 305463 | 0.00 | 19.91 | FALSE |
| MBDT | rs7125376 | 11 | 97762175 | A | G | 0.15 | 0.03 | 0.48 | 1.29E-05 | 305463 | 0.00 | 19.02 | FALSE |
| MBDT | rs72738704 | 15 | 78427490 | G | C | 0.15 | 0.04 | 0.32 | 4.05E-05 | 305463 | 0.00 | 16.85 | FALSE |
| MBDT | rs72987576 | 6 | 136944450 | C | T | -0.39 | 0.09 | 0.05 | 1.44E-05 | 305463 | 0.00 | 18.81 | FALSE |
| MBDT | rs73359181 | 8 | 140091952 | G | T | 0.23 | 0.05 | 0.12 | 6.29E-06 | 305463 | 0.00 | 20.40 | FALSE |
| MBDT | rs75516796 | 8 | 3025993 | G | A | 0.43 | 0.10 | 0.02 | 1.82E-05 | 305463 | 0.00 | 18.37 | FALSE |
| MBDT | rs76093248 | 1 | 147608966 | C | T | -0.72 | 0.17 | 0.02 | 2.98E-05 | 305463 | 0.00 | 17.43 | FALSE |
| MBDT | rs78726285 | 1 | 187694968 | C | A | -0.55 | 0.13 | 0.03 | 3.30E-05 | 305463 | 0.00 | 17.23 | FALSE |
| MBDT | rs788522 | 5 | 29475346 | G | A | 0.16 | 0.04 | 0.23 | 4.82E-05 | 305463 | 0.00 | 16.52 | FALSE |
| MBDT | rs7898239 | 10 | 10576090 | T | G | -0.24 | 0.06 | 0.13 | 1.16E-05 | 305463 | 0.00 | 19.23 | FALSE |
| MBDT | rs7910841 | 10 | 33125010 | A | G | 0.16 | 0.04 | 0.28 | 2.53E-05 | 305463 | 0.00 | 17.74 | FALSE |

| Trait | SNP | Chr | Position | EA | OA | Beta | Se | EAF | *P*-value | Samplesize | $R^2$ | F | Outlier SNPs |
|---|---|---|---|---|---|---|---|---|---|---|---|---|---|
| MBDT | rs940335 | 7 | 99561710 | G | C | 0.27 | 0.06 | 0.08 | 7.01E-06 | 305463 | 0.00 | 20.19 | FALSE |
| MBDT | rs9479979 | 6 | 154923117 | G | A | 0.18 | 0.04 | 0.20 | 4.38E-05 | 305463 | 0.00 | 16.70 | FALSE |
| MBDT | rs973679 | 19 | 46558307 | C | T | 0.18 | 0.04 | 0.77 | 2.82E-05 | 305463 | 0.00 | 17.53 | FALSE |
| MBDT | rs9928685 | 16 | 79846075 | T | A | -0.16 | 0.04 | 0.30 | 4.88E-05 | 305463 | 0.00 | 16.49 | FALSE |

Note: MBDT: mental and behavioral disorders due to the use of tobacco; SNP: single nucleotide polymorphism; Chr: chromosome; EA: effect allele; OA: other allele; Se: standard error; EAF: effect allele frequency.

**Supplementary Table 7** The results of the pleiotropy test.

| Exposure | Outcome | Methods | Egger_intercept | PRESSO_RSS | *P*-value |
|---|---|---|---|---|---|
| Dietary fibre intake[a] | MBDT | MR Egger intercept test | 0.01 | - | 0.416 |
| | | MR PRESSO global test | - | 127.77 | 0.053 |
| Dietary fibre intake[b] | MBDT | MR Egger intercept test | 0.01 | - | 0.618 |
| | | MR PRESSO global test | - | 73.24 | 0.947 |

Note: [a]: before removing outlier SNPs; [b]: after removing outlier SNPs. RSS: residual sum of squares; MBDT: mental and behavioral disorders due to the use of tobacco.

**Supplementary Table 8** The results of the heterogeneity test

| Exposure | Outcome | Methods | Q | Q_df | *P*-value |
|---|---|---|---|---|---|
| Dietary fibre intake[a] | MBDT | MR Egger | 124.32 | 100 | 0.050 |
| | | IVW | 125.15 | 101 | 0.052 |
| Dietary fibre intake[b] | MBDT | MR Egger | 71.54 | 92 | 0.944 |
| | | IVW | 71.79 | 93 | 0.950 |

Note: [a]: before removing outlier SNPs; [b]: after removing outlier SNPs. IVW: inverse variance weighted; MBDT: mental and behavioral disorders due to the use of tobacco.

**Supplementary Table 9** MR analysis results after excluding outlier SNPs.

| Exposure | Outcome | Method | nSNP | OR | 95%CI/CrI | *P*-value |
|---|---|---|---|---|---|---|
| | | MR Egger | 94 | 0.59 | (0.27-1.29) | 0.188 |
| | | WM | 94 | 0.71 | (0.45-1.11) | 0.129 |
| Dietary fibre intake | MBDT | IVW | 94 | 0.71 | (0.52-0.97 | **0.029** |
| | | PRESSO | 94 | 0.71 | (0.54-0.93) | **0.015** |
| | | BWMR | 94 | 0.69 | (0.50-0.95) | **0.024** |

Note: MBDT: mental and behavioral disorders due to the use of tobacco; IVW: inverse variance weighted; WM: Weighted median; BWMR: Bayesian weighted Mendelian randomization; SNP: single-nucleotide polymorphisms; OR: odds ratio; CI: confidence interval (reported for MR-Egger, WM, IVW, and PRESSO); CrI: credible interval (reported for BWMR). Results with statistical significance are displayed in bold.

**Supplementary Table 10** The results of the reverse MR analysis.

| Exposure | Outcome | Method | nSNP | OR | 95%CI | *P*-value |
|---|---|---|---|---|---|---|
| | | MR Egger | 89 | 1.00 | (0.99-1.01) | 0.690 |
| | | WM | 89 | 0.99 | (0.98-1.00) | 0.241 |
| MBDT[a] | Dietary fibre intake | IVW | 89 | 1.00 | (0.99-1.01) | 0.975 |
| | | PRESSO | 89 | 1.00 | (0.99-1.01) | 0.975 |
| | | BWMR | 89 | 1.00 | (1.00-1.01) | 0.474 |
| | | MR Egger | 85 | 1.00 | (0.99-1.01) | 0.851 |
| | | WM | 85 | 0.99 | (0.98-1.00) | 0.242 |
| MBDT[a] | Dietary fibre intake | IVW | 85 | 1.00 | (0.99-1.01) | 0.887 |
| | | PRESSO | 85 | 1.00 | (0.99-1.01) | 0.874 |

| Exposure | Outcome | Method | nSNP | OR | 95%CI | *P*-value |
|---|---|---|---|---|---|---|
| | | BWMR | 85 | 1.00 | (0.99-1.01) | 0.575 |

Note: [a]: before removing outlier SNPs; [b]: after removing outlier SNPs. MBDT: mental and behavioral disorders due to the use of tobacco; IVW: inverse variance weighted; WM: Weighted median; BWMR: Bayesian weighted Mendelian randomization; SNP: single-nucleotide polymorphisms; OR: odds ratio; CI: confidence interval (reported for MR-Egger, WM, IVW, and PRESSO); CrI: credible interval (reported for BWMR).

**Supplementary Table 11** The results of pleiotropy analysis in the reverse MR

| Exposure | Outcome | Methods | Egger_intercept | PRESSO_RSS | *P*-value |
|---|---|---|---|---|---|
| MBDT | Dietary fibre intake[a] | MR Egger intercept test | 0.00 | - | 0.608 |
| | | MR PRESSO global test | - | 91.04 | 0.447 |
| MBDT | Dietary fibre intake[b] | MR Egger intercept test | 0.00 | - | 0.900 |
| | | MR PRESSO global test | - | 68.73 | 0.907 |

Note: [a]: before removing outlier SNPs; [b]: after removing outlier SNPs. MBDT: mental and behavioral disorders due to the use of tobacco; RSS: residual sum of squares.

**Supplementary Table 12** The results of heterogeneity analysis in the reverse MR

| Exposure | Outcome | Methods | Q | Q_df | *P*-value |
|---|---|---|---|---|---|
| MBDT | Dietary fibre intake[a] | MR Egger | 88.70 | 87 | 0.429 |
| | | IVW | 88.97 | 88 | 0.451 |
| MBDT | Dietary fibre intake[b] | MR Egger | 67.23 | 83 | 0.896 |
| | | IVW | 67.24 | 84 | 0.910 |

Note: [a]: before removing outlier SNPs; [b]: after removing outlier SNPs. MBDT: mental and behavioral disorders due to the use of tobacco; IVW: inverse variance weighted.
